# MetaFemina: development and evaluation of a large language model-assisted platform for automated meta-analysis of nutritional exposures and breast, ovarian, and uterine cancer risk

**DOI:** 10.64898/2026.08.18.26360713

**Authors:** Margaux Delporte, Rulla Tamimi, Saurabh Mehta, Eunji Choi, Yongxian Zhang, Yushu Shi

## Abstract

**Objective:** To develop and evaluate an automated large language model (LLM)-based framework for conducting meta-analyses of nutrition-related exposures and the risk of breast, ovarian, and uterine cancers.

**Design:** We developed MetaFemina, an automated evidence-synthesis pipeline for women’s cancers that integrates keyword-based literature retrieval, LLM-assisted evidence extraction, and random-effects meta-analysis. We evaluated its performance against two recently published peer-reviewed meta-analyses and compared exposure–outcome associations across the three cancer types.

**Data sources:** PubMed articles identified through keyword-based searches of titles and abstracts.

**Methods:** MetaFemina was developed as a web platform that identifies relevant scientific articles, automatically extracts relevant information using LLMs, and synthesizes extracted evidence using random-effects meta-analysis. Additional analyses included assessment of heterogeneity, publication bias, and leave-one-out sensitivity analyses. The platform also provides sample size calculations based on synthesized effect sizes and generates visual summaries and plain-language interpretations.

**Results:** Compared with two recent peer-reviewed meta-analyses of folate and vitamin E intake in relation to breast cancer risk, MetaFemina demonstrated high sensitivity (81.82% and 80% respectively) in identifying eligible studies and additionally retrieved relevant articles that had been missed by manual screening (27 and 13 respectively). Among 226 exposures considered, lutein and *β*-carotene were significantly associated with lower risks of breast, ovarian, and uterine cancers. Vitamin D, antioxidants, and soy were significantly associated with lower risks of both breast and ovarian cancers, whereas calcium and folic acid were significantly associated with lower risks of both breast and uterine cancers. In contrast, iron, red meat, and copper were significantly associated with higher risks of both breast and uterine cancers. *ω*-6 fatty acids showed contrasting associations, being significantly associated with higher breast cancer risk but lower ovarian cancer risk. After restriction to dietary-intake studies, these cross-cancer significant associations remained statistically significant except for copper, which no longer met the two-study threshold for either breast or uterine cancer. Additionally, calcium became significantly associated with lower ovarian cancer risk, resulting in significant negative associations across all three cancer types, while vitamin E became significantly associated with lower breast cancer risk and remained significantly associated with lower ovarian cancer risk.

**Conclusions:** MetaFemina demonstrated high sensitivity for identifying relevant scientific literature, extracts key evidence, and performs statistically rigorous automated meta-analyses. The framework may facilitate more rapid evidence synthesis in nutritional epidemiology and may support researchers in study design, hypothesis generation, and interpretation of emerging evidence.

## 1 Introduction

Our society is facing an ever-expanding volume of health information. Yet, much of the health information directed at the general public is not grounded in rigorously peer-reviewed research and is often based on isolated findings rather than conclusions that have been consistently evaluated across multiple studies [1]. Meanwhile, nutrition scientists, epidemiologists, clinicians, and the general public are actively seeking nutritional interventions that may reduce disease risk [2]. Breast cancer, a hormone-related malignancy, is the most commonly diagnosed cancer among women in the United States. Prior studies have implicated obesity [3], hormonal pathways [4], the immune system [5], and chronic inflammation [6] in breast cancer development, and many of these factors are closely linked to modifiable dietary behaviors. In a narrative review, Rodŕıguez et al. [7] reported that breast, uterine, and ovarian cancers share many epidemiological, lifestyle, and local hormonal and metabolic factors. A comprehensive synthesis of the published literature would help identify promising dietary interventions and facilitate future study designs by providing sample size calculations based on meta-analytic evidence.

Rigorous evidence synthesis provides structured approaches for identifying relevant studies, appraising study quality, extracting evidence, and quantitatively summarizing findings [8]. However, manually conducting these processes is often labor-intensive and difficult to scale across the rapidly expanding biomedical literature. This limitation highlights the need for a freely accessible tool that can contextualize and evaluate the most up-to-date nutritional health claims through meta-analysis on a large scale for researchers and the general public.

Large language models (LLMs) are creating new opportunities for evidence synthesis [9]. Although early studies recommended cautious adoption because of concerns about accuracy and reliability [10, 11], more recent work has demonstrated strong potential when advanced prompting strategies are used [12]. For example, SciDaSynth [13] has demonstrated effectiveness in producing high-quality structured data more efficiently than baseline methods through a within-subjects study. Cao et al. [14] reported that LLM-based abstract and full-text screening achieved sensitivity and specificity above 85%. More recent LLM-driven workflows have also shown promise in supporting both fully automated and human-in-the-loop systematic review processes [15]. Existing tools such as Elicit[16] and TrialMind [17] primarily demonstrate the conceptual potential of using LLMs to support systematic reviews, rather than providing free, reliable, and end-to-end meta-analytic product on specific diseases.

In this paper, we present an LLM-based tool that conducts meta-analyses on peer-reviewed abstracts and full texts linking nutritional exposures with breast, ovarian, and uterine cancers. The tool automates the full pipeline of retrieval, extraction, analysis, interpretation, and diagnostic statistics, while still allowing human verification. The platform also provides sample size calculations based on synthesized effect sizes and generates visual summaries and plain-language interpretations. The website is updated monthly to incorporate the latest publications and supports multiple languages to facilitate its use by the global scientific community. In Section 2, we provide a detailed overview of the tool’s scope, as well as the methods underlying the search, extraction, and statistical analysis. In Section 3, we present two case studies examining the associations of folic acid and vitamin E with breast cancer risk and compare the findings with those of recently published peer-reviewed meta-analyses. Section 4 presents an overview of our results for all considered nutritional exposures and compares their effects across the three cancer types. Finally, Section 5 provides the discussion, and Section 6 presents the concluding remarks and future directions for MetaFemina.

## 2 Methods

Figure 1 provides an overview of the MetaFemina automated pipeline. Dark pink boxes denote external resources, light pink boxes denote analysis inputs, green boxes denote analytical procedures, and gray boxes denote generated outputs. Precomputed meta-analysis results are available at this website.Users can access MetaFemina’s synthesized evidence and summarized results without purchasing tokens, configuring external services, or having prior knowledge of large language model APIs. Additional technical details can be found in Appendix A.

**Figure 1:**
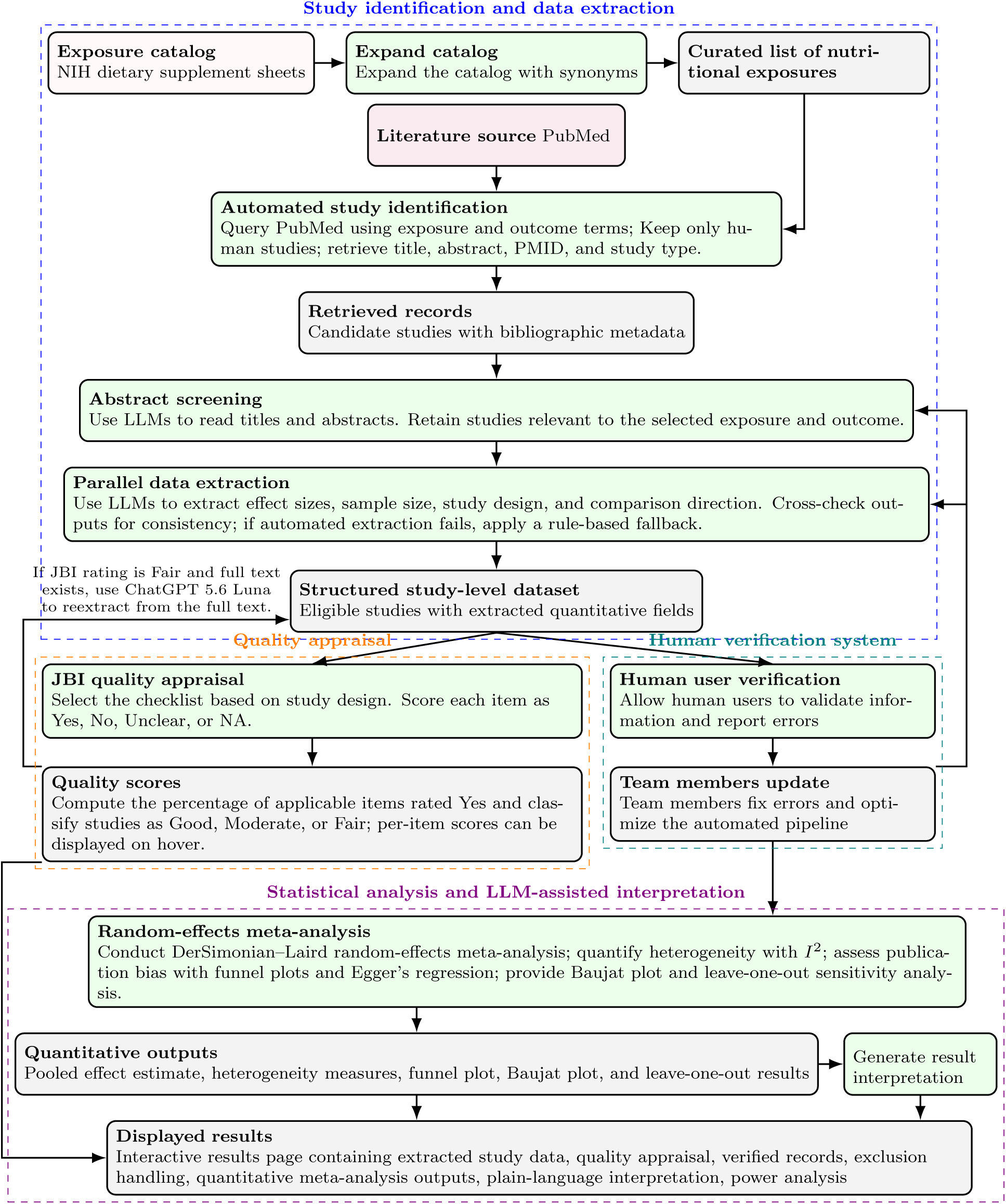
Overview of the MetaFemina automated evidence synthesis pipeline, including literature retrieval, LLM-assisted screening and extraction, quality appraisal, human verification, and meta-analysis.

### 2.1 Study Identification and Data Extraction

We compiled a list of nutrition-related exposures from the NIH dietary supplement fact sheets [18] and expanded it with synonyms to ensure that pertinent studies were captured. We then searched the PubMed database through the API to identify relevant studies. As shown in the snapshot Figure 2, MetaFemina displays the synonyms of each exposure used in each PubMed query for transparency and clarity.

**Figure 2:**
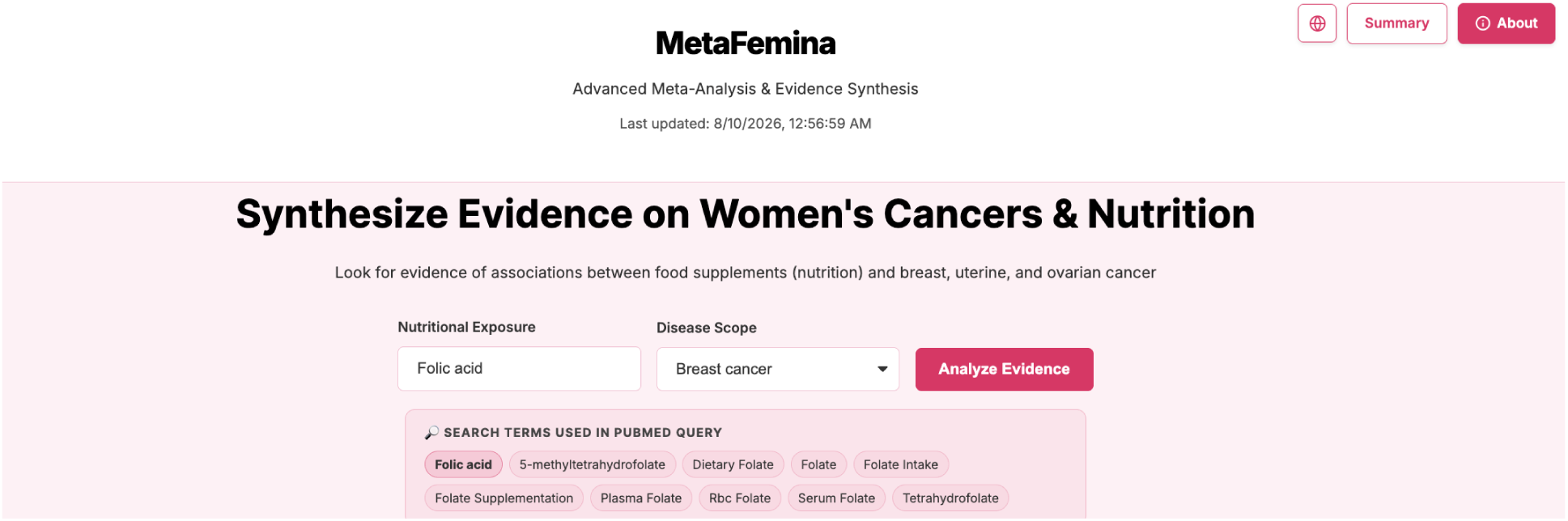
User interface of MetaFemina: selection of a nutritional exposure displays its corresponding synonyms and related terms used for literature identification.

The default setup for MetaFemina limits the search to human studies and excludes review and meta-analysis articles to avoid double-counting evidence. Further, to maintain a focus on the general population, we excluded studies that examined interactions between nutrition and specific genotypes or family history of breast or gynecological cancer. For each retrieved study, detailed study information, such as title, abstract, and study type, is stored for further analysis. More information on the literature search can be found in Appendix B.

For the retrieved studies, we first screened the abstracts to confirm relevance to the selected exposure and study type with an LLM. We then used LLMs (OpenAI GPT-4o and Google Gemini 2.5 Flash) to extract key information, including effect sizes such as relative risk (RR), hazard ratio (HR), or odds ratio (OR), sample size, study design, exposure quantification, and the comparison being made. When the number of cases was not available, we estimated the number based on the sample size and lifelong incidence estimates [19, 20, 21]. To improve reliability, the two models were run in parallel and their outputs were cross-checked for consistency, particularly with respect to the direction of association (for example, high-versus-low exposure). Overall, this pipeline enables rapid and reliable extraction of relevant information from large numbers of studies. More technical details on the data extraction can be found in Appendix C.

A challenge in both manual and automated evidence synthesis is exposure standardization: determining what qualifies as a “high” exposure. For example, if two studies each define high and low exposure using the study-specific median as the cutoff, the resulting cutoffs are likely to differ across studies. Similarly, across studies examining the same topic, one study may dichotomize the exposure into high and low groups, whereas others may categorize it into tertiles or quartiles. If we impose overly strict cutoff requirements, only a small number of studies will be considered comparable, which undermines the purpose of conducting a large-scale meta-analysis. Therefore, to preserve the broader evidence base, we synthesize studies even when exposure cutoffs are study-specific. When a study reports multiple exposure levels, such as low, medium, and high intake, we use the effect comparing the highest exposure group with the lowest exposure group, which is the most conventional approach in nutritional studies given the paywall limitation of the closed-access articles [22] and has been repeatedly adopted by previous meta-analyses [23, 24, 25, 26, 27].

Another common problem encountered even with manual evidence synthesis is the integration of dietary intake measures and biospecimen-based measurements. We acknowledge that high dietary intake is not equivalent to a high concentration measured in a biological specimen. In some cases, high intake may even reflect poor absorption or nutritional insufficiency at the individual level, and the relationship between intake and biospecimen concentration may be negative. Nevertheless, to provide a broad overview, we retain results from both dietary intake studies and biospecimen-based analyses, which is a common practice for high-versus-low group comparison adopted by previous meta-analyses [28, 29]. Specifically, dietary intake measures include exposures assessed by food frequency questionnaires, dietary recalls, diet records, supplement intake reports, self-reported intake, and dietary patterns. Biospecimen-based measures include exposures assessed from human biological specimens, such as blood, serum, plasma, urine, saliva, tissue, fecal samples, metabolomics profiles, biomarker concentrations, or similar biological measurements.

On our website, for each paper, we will display how the exposure was measured, supported by a short excerpt from the original publication. By default, all papers will be included, while users may apply filters to focus on specific exposure measurement types. Forest plots summarizing only studies based on dietary intakes are presented in the supplemental Section F.2.

### 2.2 Quality Appraisal

Study quality is assessed automatically by the LLMs using the Joanna Briggs Institute (JBI) Critical Appraisal Checklists [30]. The specific items evaluated depend on the study design: cohort studies, case-control studies, randomized controlled trials, and cross-sectional designs (fallback). The LLM evaluates each item as “Yes”, “No”, “Unclear”, or “NA” based on the abstract content. A quality percentage is calculated as the proportion of applicable items rated “Yes”, with studies categorized as Good (*>*80%), Moderate (51–80%), or Fair (*<*51%). In the MetaFemina web platform, per-item scores for each study are displayed when hovering over the quality appraisal, with additional details about the items available on the “About” page.

### 2.3 Automated Statistical Analysis

We combine results across studies using the DerSimonian–Laird random-effects meta-analysis [31], which allows the underlying effect size to vary across studies rather than assuming a single common effect. This framework remains appropriate when studies differ in patient populations, specific exposure definitions, follow-up time, or other design features [32]. As suggested by Grant [33], Moser and McCann [34], and Cherif and Dziri [35], relative risk is often more intuitive for communicating exposure effects and more suitable for summarizing evidence across studies in a meta-analysis. Therefore, to harmonize effect measures across studies, we convert studies reporting ORs or HRs to relative RRs for meta-analytic effect size calculation, assuming a baseline breast cancer incidence of *p*_0_ = 0.13 [20], an ovarian cancer incidence of *p*_0_ = 0.013 [36] and a uterine cancer incidence of *p*_0_ = 0.031 [21]. We note that often HR and OR were used as an approximation to the RR in many previous meta-analyses [37, 25, 23, 38]. However, given the high lifelong incidence of breast cancer, we conducted the following conversion for more accurate calculation. Specifically, ORs are converted using 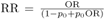, and HRs are converted under the proportional hazards assumption using RR = *{*1*−*(1 *−p*_0_)^HR^*}/p*_0_. Although RR is displayed as the default effect measure, users can also choose to display the effect sizes in HRs or ORs on the MetaFemina website. Additionally, we treated incidence rate ratio (IRR) as RR in meta-analysis calculation. Although the incidence rate ratio (IRR) compares event rates per unit person-time, whereas the risk ratio (RR) compares cumulative risks over a specified period, the two measures can provide close approximations of the same relative association, and treating IRRs as RRs in meta-analysis is consistent with conventions adopted in previous studies [39]. For each exposure, we provide a 95% confidence interval for the estimated meta-analytic effect size, as well as the 95% prediction interval for a future study. Studies reporting other types of effect measures, such as the population attributable fraction (PAF), are included in the extracted study list but are not incorporated into the meta-analysis.

We assess the consistency of results across studies using both the between-study variance *τ* ^2^ and the *I*^2^ statistic. Under a random-effects model, *τ* ^2^ quantifies the dispersion of the true study-specific effects around the average effect, and *I*^2^ quantifies the percentage of the total variability that is attributable to between-study heterogeneity rather than sampling error [40]. Higher *τ* ^2^ and *I*^2^ values indicate greater inconsistency across studies and suggest that the underlying effect sizes may differ more substantially from one study to another. We provide an automatic interpretation of *I*^2^ based on guidance from the Cochrane Handbook [41]. Specifically, *I*^2^ values of 0%–40%, 30%–60%, 50%–75%, and 75%–100% are interpreted, respectively, as “heterogeneity that might not be important”, “moderate heterogeneity”, “substantial heterogeneity”, and “considerable heterogeneity”.

Potential publication bias is assessed using funnel plots and Egger’s regression test [42]. If the number of studies is fewer than 10 for an exposure, the Egger’s test will not be conducted. Funnel plots display individual study log effect sizes against their standard errors, with pseudo-95% confidence limits around the pooled estimate. Egger’s test regresses the standardized effect size on study precision; a statistically significant intercept suggests potential asymmetry. Alongside these diagnostics, we provide an automatically generated narrative interpretation of the funnel plot and Egger’s test results. As sensitivity analyses, we further provide Baujat plots [43] and leave-one-out analyses to identify influential studies and assess the robustness of the pooled estimate. More details on the statistical approach can be found in Appendix D.

### 2.4 Inclusion & Exclusion Options, Verification System and LLM Interpretation

MetaFemina focuses on primary women’s cancer incidence rather than metastatic cancers. Because the focus of the web platform is on women’s cancers and potentially different physiological mechanisms, MetaFemina excludes papers on male breast cancer. Although MetaFemina is designed as an automated evidence synthesis system, it gives biomedical researchers full control beyond the default settings. For each included study, we display key study-level information, including the first author’s last name, journal name, original effect measure, and effect size as reported in the study. Note that, for harmonized random-effects meta-analysis, these effect estimates may be converted to relative risks. The converted relative risk is displayed in light gray text below the original effect size reported in the study.

Users can sort studies by the last name of the first author, publication year, sample size, journal name, JBI rating, and effect size to quickly identify studies of interest. We also provide four key filters to support customized evidence review: study design (whether the study is prospective, retrospective, or a clinical trial), minimum number of cases, study quality, and reported effect measure (relative risks, odds ratios, or hazard ratios). By default, MetaFemina sets the minimum number of cases to 50, restricting the primary analysis to moderate-to-large studies, and includes only studies rated as moderate quality or above using the JBI score [30]. Studies with the number of cases or JBI quality score below these thresholds are excluded from the meta-analysis. In the website interface, these studies remain visible but are visually de-emphasized using a gray overlay to indicate that they are not included in the current analysis. However, users may manually adjust the inclusion threshold or select specific studies for inclusion. The information of the selected studies can be exported as a CSV file by clicking on the pink “Export” button.

MetaFemina adopts a crowdsourced verification mechanism. Studies receiving multiple independent validations are considered to have greater support for inclusion. Conversely, users may flag a study if they identify errors in the extracted information or believe that the study does not meet the inclusion criteria. Once a study receives two flags, an automated email notification is sent to the research team for manual review. For each study, the website displays the number of times it has been verified or excluded by human users. Repeated verification or exclusion by independent users provides additional contextual information about the study’s quality and relevance to the topic under investigation.

Two automatically generated narrative summaries explain the results in plain language: one interprets the pooled meta-analysis estimate and heterogeneity metrics, as illustrated in Figure 5; the other interprets the funnel plot and publication-bias assessment, as illustrated in Figure 6.

## 3 Comparison with published meta-analyses

In this section, we compare MetaFemina with two recently published, peer-reviewed meta-analyses. The exposures, folic acid and vitamin E, were selected based on the recency of the corresponding publications rather than their status as established or emerging risk factors.

### 3.1 Example 1: Folic acid intake and breast cancer risk

As a first illustrative example, we will use MetaFemina to examine the association between folic acid intake and breast cancer risk and compare the results with those from a recent peer-reviewed meta-analysis on dietary folic acid intake [44], published in 2025. Through this comparison, we provide a step-by-step overview of MetaFemina.

To start, the user selects a nutritional exposure from the dropdown menu (in this example, folic acid is selected). Upon selection, a list of synonyms and relevant terms is displayed in a pink box (Figure 2). To improve the coverage of relevant literature, MetaFemina groups conceptually related exposures when querying the NCBI database. We recognize that folic acid is the synthetic form of vitamin B9, whereas folate is its naturally occurring form, and that these compounds may undergo different metabolic pathways in the human body. Nevertheless, to provide a comprehensive overview of the association between vitamin B9 and breast cancer risk, we include studies evaluating either folic acid or folate. This approach is also adopted by the comparator meta-analysis [44]. Also, consistent with the comparator meta-analysis [44], MetaFemina encompasses studies on dietary folate, a combination of dietary folate with folic acid from fortified foods and supplements, and supplemental folic acid only.

As the first step, an automatically generated query is searched in PubMed using the PubMed API (See Supplementary Materials B). For this exposure-outcome combination, the pipeline initially identified 866 studies, which were then sent to a screening layer that uses an LLM to verify if the study is directly associated with the exposure. If a study is excluded, its PMID, the reason for exclusion, exposure, and disease scope will be recorded. Once the search, screening, and extraction are completed, a forest plot is automatically generated (Figure 3). This forest plot can be dynamically updated based on the studies selected by the user from the Extracted Studies Table (Figure 4). Users can select specific studies and update the meta-analysis to include only the selected studies by clicking the “Update analysis” button. In the Extracted Studies Table, MetaFemina provides the key metadata for each study used to determine inclusion or exclusion, the quantitative data used in the meta-analysis, and additional descriptive information, including the evaluated comparison, study population, study design, and excerpt from the original article supporting the extraction. Each study title links to its PubMed record, where the abstract and full text can be accessed. Performing meta-analysis on folic acid and breast cancer risk with OpenAI GPT-4o and Google Gemini 2.5 Flash requires approximately 1 hour and 2 minutes and incurs a cost of $5.99. Saved results can be loaded by users and are available immediately.

**Figure 3:**
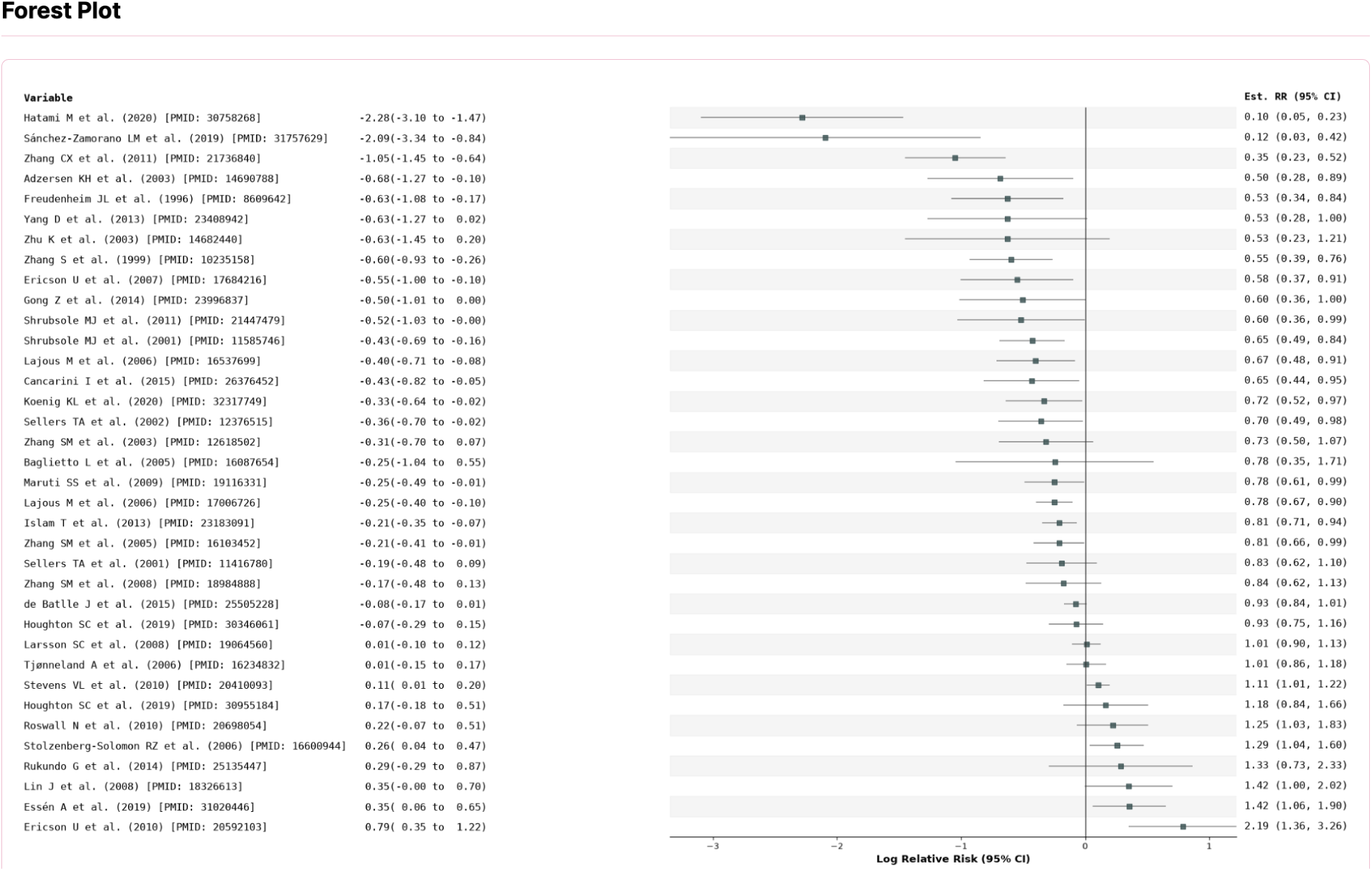
Forest plot showing individual study estimates with corresponding confidence intervals.

**Figure 4:**
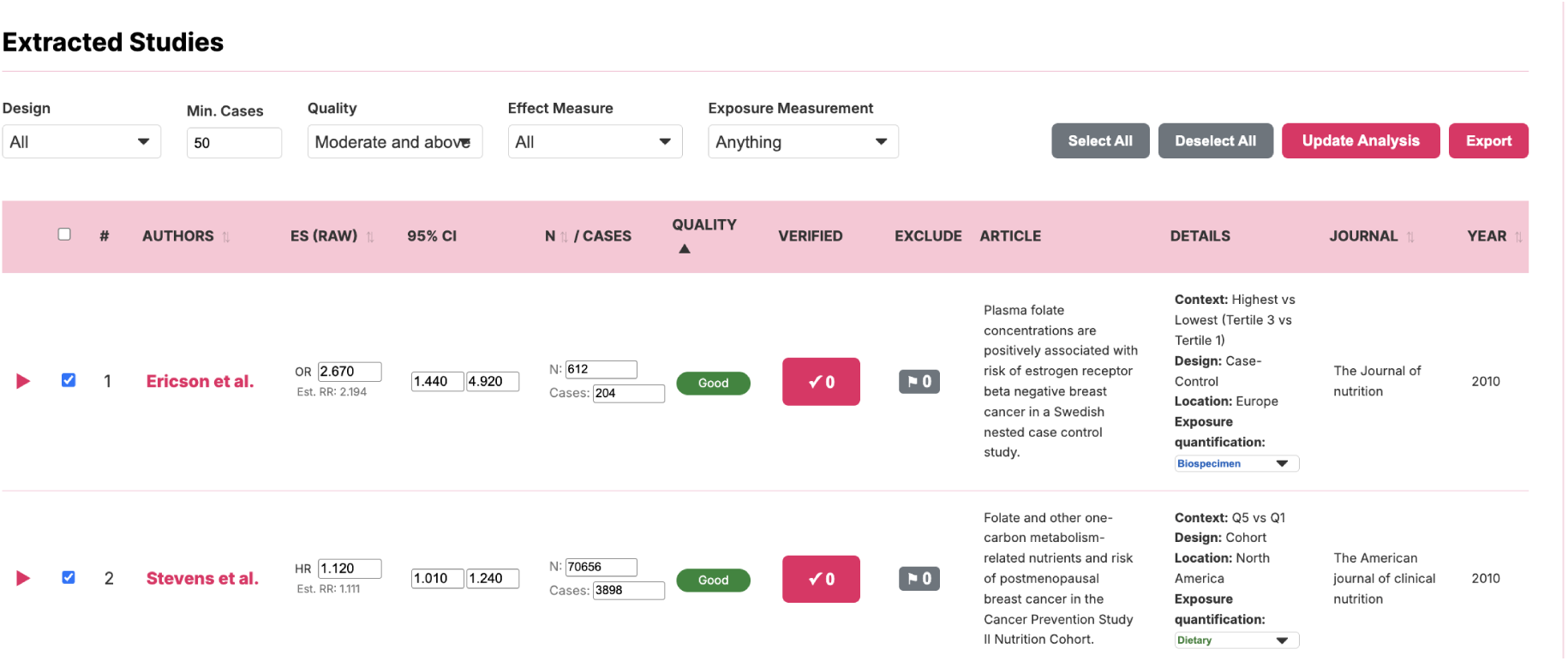
Extracted studies table displaying study-level characteristics and effect estimates of the association between folic acid and breast cancer. Users can select studies and apply filters based on study design, sample size, quality, and reported effect measure to refine the meta-analysis.

Based on the studies selected in the Extracted Studies Table, MetaFemina computes a random-effects meta-analysis (Figure 5). Plain-language interpretations of the statistical analysis, including the random-effects meta-analysis results and the heterogeneity measure *I*^2^, are generated to improve understanding and accessibility for broad audiences. The specific wording is tailored to the effect-size measure and the corresponding test results.

**Figure 5:**
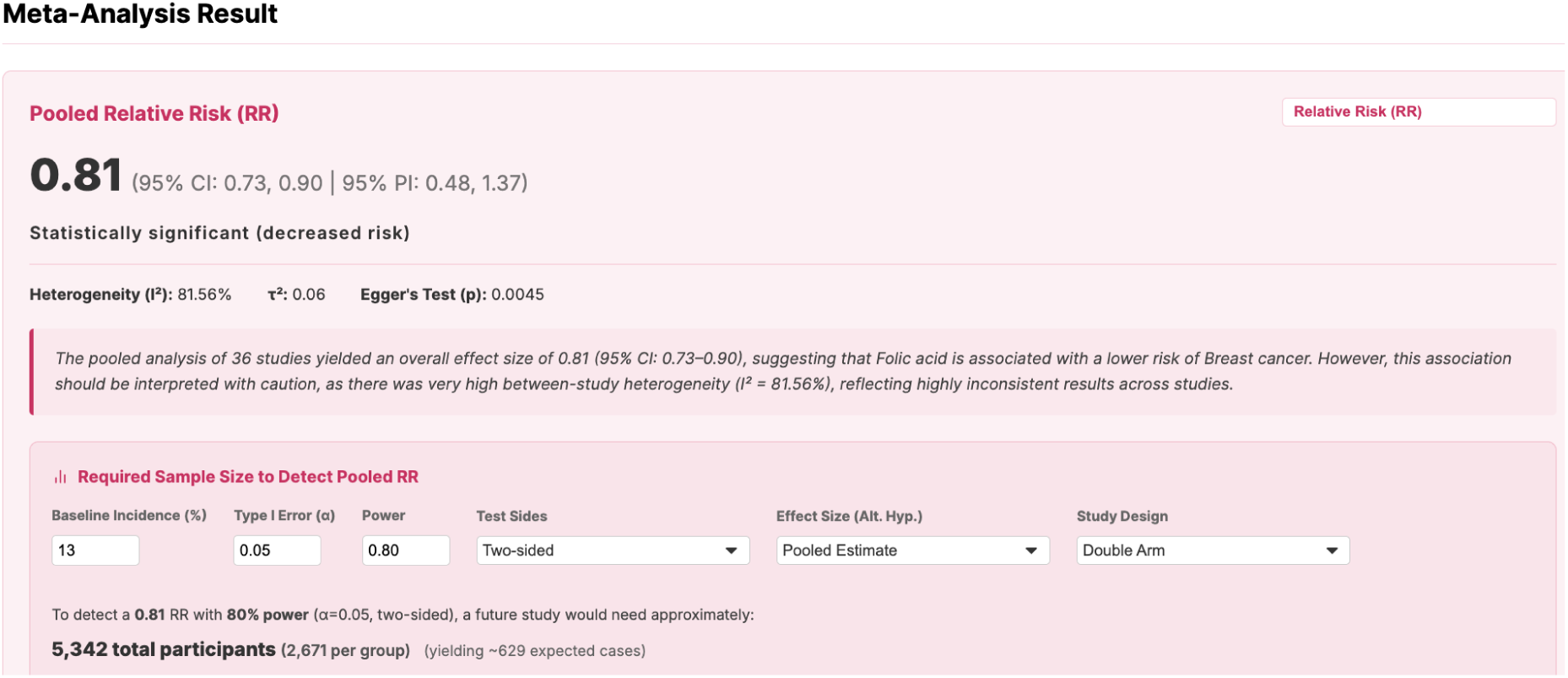
Pooled effect size with confidence and prediction intervals, heterogeneity assessment, and automatically generated plain-language interpretation of the association between folic acid and breast cancer incidence. Additionally, a simple power analysis tool is provided.

For the broad category of folic acid–related exposures and breast cancer, the pooled analysis of 36 studies yielded an estimate of 0.81 (0.73–0.90), indicating an association with a lower risk of breast cancer. However, this finding should be interpreted cautiously because between-study heterogeneity was considerable (*I*^2^ = 81.56%), indicating considerable inconsistency across studies. In addition, Egger’s test was significant (*p* = 0.0045), suggesting possible small-study effects, including publication bias. When the analysis was restricted to 28 studies based only on dietary intake, the pooled estimate decreased further to 0.74 (0.66– 0.83). Nevertheless, considerable heterogeneity and evidence of possible small-study effects remained, with *I*^2^ = 82.09% and Egger’s test *p <* 10*^−^*^4^.

Because one of the primary applications of our tool is to support researchers in designing new studies, we incorporated a sample size calculator for prospective studies comparing breast, ovarian, or uterine cancer risk between individuals with high versus low nutritional exposure. The sample size calculator uses the meta-analytic relative risk as the alternative hypothesis, while allowing users to make more or less conservative assumptions by using the lower or upper bound of the confidence interval or prediction interval. Relevant parameters can be adjusted by the researcher, including the baseline incidence rate (default: 13% for breast cancer, 3.1% for uterine cancer, and 1.3% for ovarian cancer); the significance level *α* (default 0.05); power (default 0.8); one-sided versus two-sided testing (default two-sided); and single-arm versus two-arm designs (default two-arm). As shown in Figure 5, using the updated pooled estimate of 0.81 for folic acid and breast cancer risk, a future study would require approximately 5,342 total participants (2,671 per group). This sample size is expected to yield approximately 629 cases.

Next, publication bias is assessed using a funnel plot and Egger’s test, accompanied by an automatically generated plain-language interpretation of the findings (Figure 6). For the folic acid and breast cancer risk example, Egger’s test indicated significant funnel plot asymmetry (*p* = 0.0045), suggesting potential small-study effects, including publication bias. Because funnel plot asymmetry can arise from factors other than publication bias, this finding should not be interpreted as definitive evidence of selective publication. Nevertheless, such asymmetry may affect the reliability of the pooled estimate, and sensitivity analyses should be considered to assess the robustness of the association. We also provide additional diagnostics: a Baujat plot is presented to visualize each study’s contribution to both heterogeneity and the overall effect size, alongside a leave-one-out sensitivity analysis (Figure 7).

**Figure 6:**
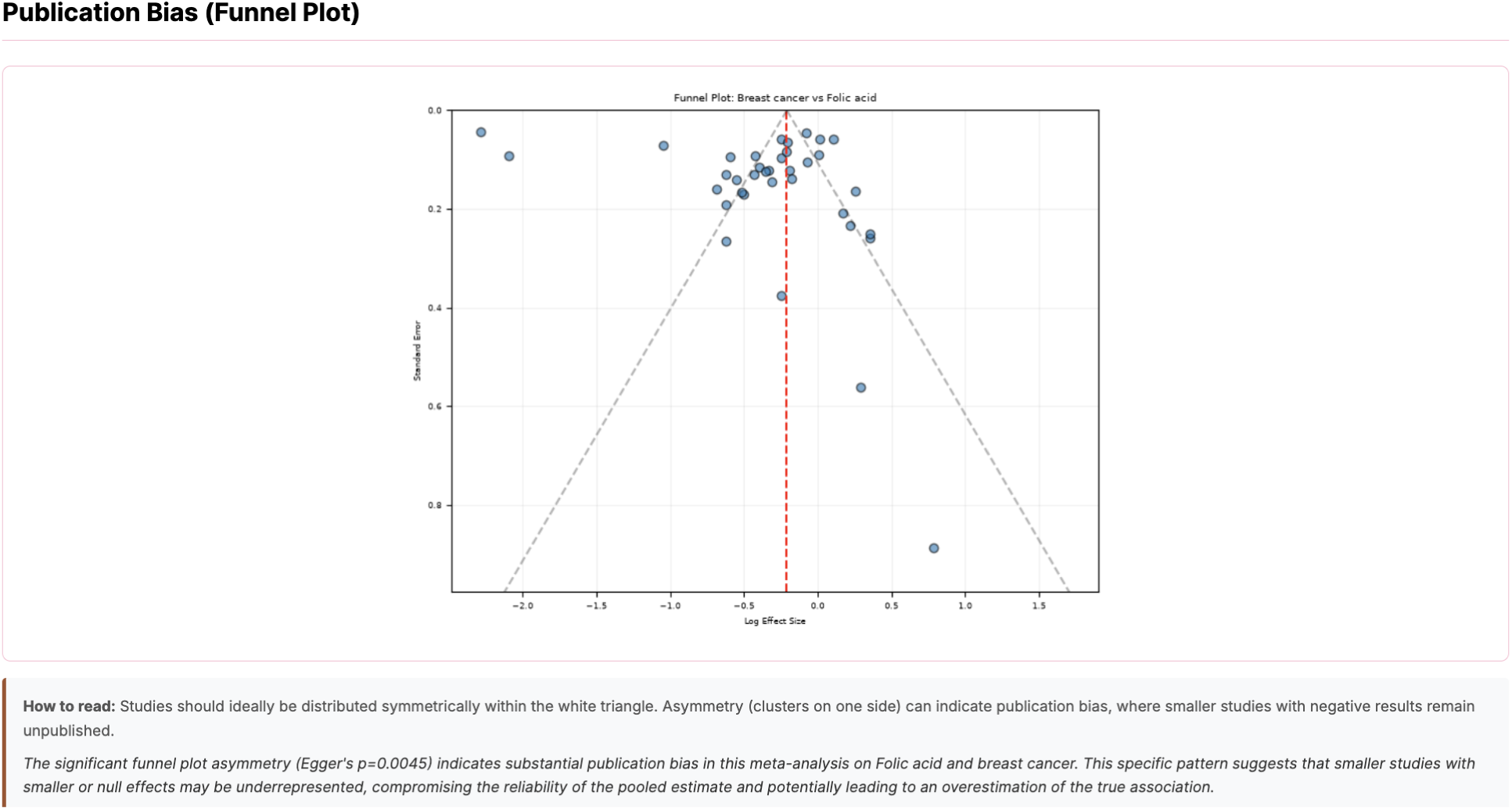
Assessment of publication bias using a funnel plot and Egger’s test of the association between folic acid and breast cancer, including auto-generated text summarizing the result in plain language.

**Figure 7:**
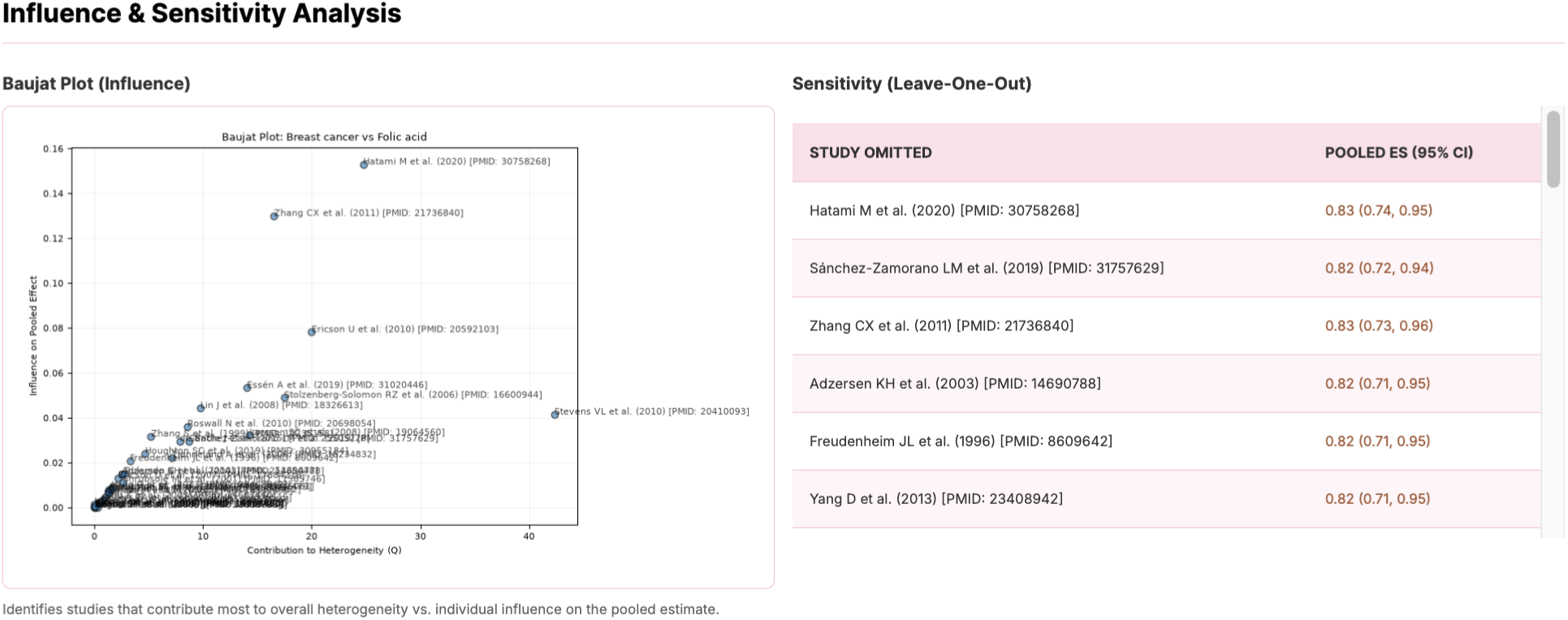
Influence and sensitivity analyses evaluating the robustness of the meta-analysis results of the association between folic acid and breast cancer to individual studies and model assumptions.

Comparing our results with those reported by Eleotério et al. [44], our tool identified 10 of the 11 studies included in their meta-analysis, attaining a sensitivity of 81.82%. Of these 10 studies, one was excluded from our meta-analysis because it included fewer than 50 cases [45]. All studies identified by Eleotério et al. [44] are shown in Table 1. The study by Van Puyvelde et al. [46], which was not included in MetaFemina and is shaded gray in Table 1, was successfully retrieved and identified as relevant by our meta-analytic tool. However, it was excluded by the automated pipeline because the exposure effect was modeled nonlinearly and no directly extractable effect-size estimate was reported. We included 27 additional relevant studies on folate intake and breast cancer risk, which are listed in Supplemental Table S1. All 27 articles were published before 2021. Light-blue rows indicate records whose exposure measurement was not classified as dietary intake by MetaFemina, including human-biospecimen-based studies, unclear exposure types, and mixed exposure quantifications. As an experiment, we asked the commercial tool Elicit [16] to conduct a meta-analysis of folic acid and breast cancer incidence. Its response summarized existing meta-analyses and outlined an appropriate review protocol, rather than fulfilling the requested task of conducting a complete systematic review and meta-analysis. In contrast, MetaFemina identifies individual studies, applies prespecified eligibility criteria, extracts and harmonizes effect estimates, performs quantitative meta-analysis, and generates outputs freely accessible to the scientific community.

**Table 1:** Studies on folate-related exposures and breast cancer risk included in Eleotério et al. [44].

| Authors | Location | Sample size/Cases | Design | Exposure and assessment | How the exposure groups or contrast were formed | Effect estimate (95% CI) |
| --- | --- | --- | --- | --- | --- | --- |
| Thorand et al. [45] | Germany | 149/43 | Retrospective case-control | 201-item FFQ; total dietary folate/folate equivalents and methionine | Low, intermediate, and high methyl-group availability; shown estimate compares high with low total folate after energy adjustment | OR 0.79 (0.51–1.21) |
| Zhang et al. [47] | USA | 88,818/3,483 | Prospective cohort | FFQ; total folate from food and supplements | $\geq 600$ versus 150–299 $\mu\text{g/day}$ of total folate | RR 0.55 (0.39–0.76) |
| Shrubsole et al. [48] | China | 2,703/1,321 | Retrospective case-control | 76-item FFQ; dietary folate | Highest versus lowest quintile of dietary folate | OR 0.62 (0.46–0.82) |
| Zhu et al. [49] | USA | 609/304 | Retrospective case-control | 192-item FFQ; methyl-group dietary intake including folate | High versus low methyl-group intake within an ERα-gene-methylation analysis; estimate reoriented from low versus high | OR 0.50 (0.2083–1.25) |
| Tjønneland et al. [50] | Denmark | 24,697/388 | Prospective cohort analysis | 192-item FFQ; dietary folate | Folate intake $> 400 \mu\text{g/day}$ vs $\leq 400 \mu\text{g/day}$ | IRR 1.01 (0.85–1.20) |
| Lajous et al. [51] | France | 62,739/1,812 | Prospective cohort | 208-item FFQ; dietary folate | Highest versus lowest quintile (Q5 versus Q1); median dietary folate 522 versus 296 $\mu\text{g/day}$ | RR 0.78 (0.67–0.90) |
| Larsson et al. [52] | Sweden | 61,433/2,952 | Prospective cohort | FFQ; dietary folate | $\geq 277$ versus $< 200 \mu\text{g/day}$ of dietary folate | RR 1.01 (0.90–1.13) |
| Maruti et al. [53] | USA | 35,023/743 | Prospective cohort | 120-item FFQ plus supplement history; total folate expressed as DFE | $\geq 1272$ versus $\leq 345$ DFE/day using approximately 10-year average intake | RR 0.78 (0.61–0.99) |
| Kim et al. [54] | Canada | 400/129 | Retrospective case-control | Supplement questionnaire; supplemental folic acid | Primary estimate: ever versus never use; dose groups were never, moderate (8.56–89.29 $\mu\text{g/day}$), and high ($> 89.29 \mu\text{g/day}$) | OR 0.45 (0.25–0.79) |
| Van Puyvelde et al. [46] | Europe | 318,686/13,320 | Prospective EPIC cohort | Country- or center-specific validated dietary questionnaires; dietary folate | Nonlinear dose-response model with approximately 205 $\mu\text{g/day}$ as the reference; risk decreased up to about 350 $\mu\text{g/day}$ before rising | Does not apply |
| Ericson et al. [55] | Sweden | 11,699/392 | Prospective cohort | Diet-history method combining FFQ, food record, and interview; dietary/total folate | Highest versus lowest quintile; median dietary folate 302 versus 153 $\mu\text{g/day}$ | HR 0.56 (0.35–0.90) |

### 3.2 Example 2: Vitamin E and breast cancer risk

As a second benchmark, we compared the performance of MetaFemina with a recently published meta-analysis investigating vitamin E supplementation and dietary intake in relation to breast cancer risk [39]. The published review incorporated five observational studies and reported no significant association between total vitamin E intake and breast cancer risk. MetaFemina retrieved substantially more studies, including recently published articles and studies not captured by the original search strategy. The only study identified by de Oliveira et al. [39] but not included in MetaFemina was excluded because its abstract reported neither an effect estimate, a confidence interval, nor a *p*-value specifically associated with vitamin E. The exclusion is consistent with our protocol described in Supplemental Section C. This study is shaded gray in Table 2. The 13 additional studies identified by MetaFemina are presented in Supplementary Table S2. For the broad category of vitamin E exposures and breast cancer, the pooled analysis of 17 studies with at least 50 cases yielded an estimate of 0.85 (0.71–1.01), indicating no statistically significant association with lower breast cancer risk. There was considerable heterogeneity among these studies (*I*^2^ = 81.77%), whereas Egger’s test did not provide evidence of small-study effects at the conventional significance threshold (*p* = 0.1288). When the analysis was restricted to dietary-intake studies, the number of included studies decreased to 12, and the pooled estimate decreased further to 0.81 (0.70–0.94), becoming statistically significant. Substantial heterogeneity remained (*I*^2^ = 64.88%), and Egger’s test was significant (*p* = 0.0011), suggesting possible small-study effects in the dietary-intake-only analysis.

**Table 2:**
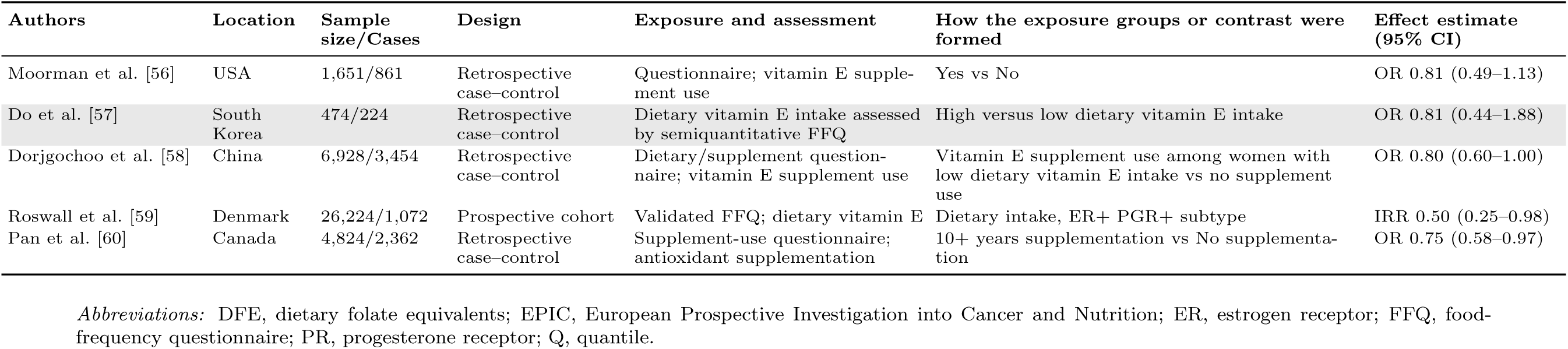
Studies on vitamin E exposures and breast cancer risk included in de Oliveira et al. [39].

## 4 Comparative analysis

We summarized exposures associated with increased and decreased breast, uterine, and ovarian cancer risk. In the summary forest plots across exposures, only exposures with a pooled effect estimate higher/lower than 1.00 and evaluated in at least two independent studies are shown; statistical significance was not required. Only studies with at least 50 cases were included. Case counts were either extracted directly from the abstract or, when unavailable, estimated from the reported total sample size. The current MetaFemina includes 736 breast cancer study records, 103 ovarian cancer study records, and 70 uterine cancer study records.

### 4.1 Exposures associated with breast cancer

For the association between exposures and breast cancer risk, the pattern of results was notably asymmetric. Among exposures evaluated in at least two studies, pooled estimates were below 1.00 for substantially more exposures than they were above 1.00, and statistically significant negative associations were also more numerous than statistically significant positive associations.

Among exposures evaluated in at least two studies and with pooled estimates above 1.00 (Figure 8), most pooled effect estimates were modest and frequently close to the null. Here we use different colors to denote various exposure types, and the same color convention is later used in heterogeneity assessment plots. Red square outlines highlight statistically significant results, and dashed lines indicate prediction intervals. Chromium 1.13 (1.06–1.21), iron 1.15 (1.05–1.25), alcohol 1.27 (1.24–1.29), red meat 1.27 (1.20–1.34), dehydroepiandrosterone 1.29 (1.17–1.41), copper 1.66 (1.07–2.56), and *ω*-6 fatty acids 1.74 (1.26–2.42) were significantly positively associated with breast cancer risk, whereas several others, including leucine 1.01 (0.91–1.13), tea 1.01 (0.92–1.11), magnesium 1.01 (0.80–1.29), carnitine 1.05 (0.83–1.35), conjugated linoleic acid 1.06 (0.92–1.22), glutamine 1.14 (0.80–1.63), zinc 1.14 (0.68–1.91), and eggs 1.19 (0.99–1.43), had positive point estimates but were not statistically significant. After restricting the analysis to dietary-intake studies, iron 1.22 (1.04–1.43), alcohol 1.24 (1.22–1.27), red meat 1.33 (1.24–1.42), and *ω*-6 fatty acids 1.74 (1.26–2.42) remained significantly positively associated with breast cancer risk.

**Figure 8:**
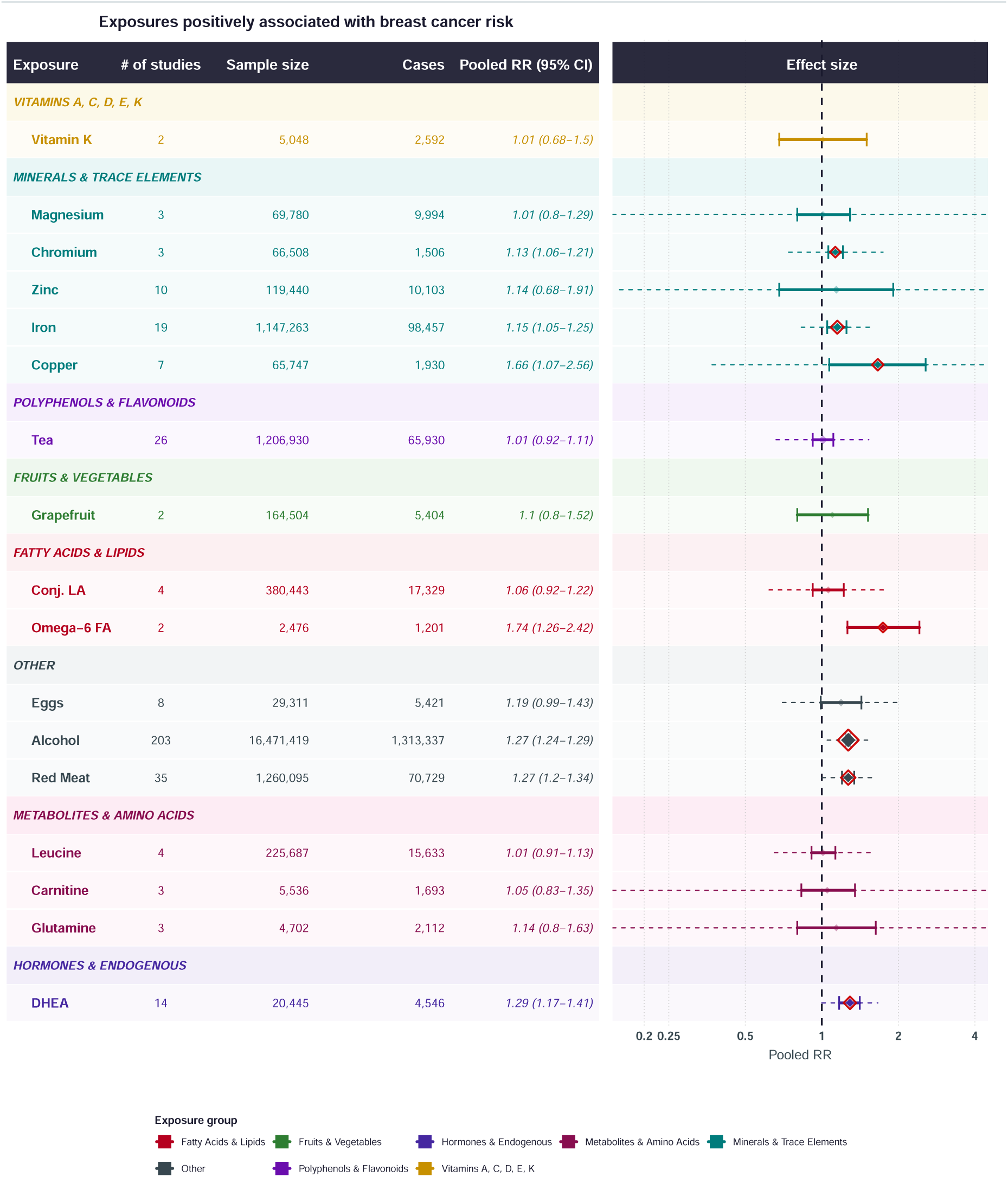
Summary forest plot of nutritional exposures with pooled estimates above 1.00 for breast cancer risk.

In contrast, the negative-association profile was broader and more pronounced (Figure 9). Significant negative associations were observed for molybdenum 0.42 (0.24–0.76), vitamin B6 0.56 (0.42–0.76), lutein 0.62 (0.52–0.74), antioxidants 0.65 (0.53–0.80), *ω*-3 fatty acids 0.65 (0.56–0.76), lycopene 0.68 (0.57–0.81), vitamin B12 0.68 (0.48–0.95), garlic 0.71 (0.55–0.94), *β*-carotene 0.73 (0.66–0.80), legumes 0.74 (0.65–0.85), olive 0.75 (0.64–0.87), soy 0.75 (0.65–0.86), vitamin B1 0.75 (0.61–0.93), Mediterranean diet 0.76 (0.70–0.83), cod liver oil 0.78 (0.68–0.90), folic acid 0.81 (0.73–0.90), vitamin D 0.82 (0.79–0.86), yogurt 0.82 (0.73–0.94), and calcium 0.91 (0.86–0.96). Several of these significant associations were supported by relatively large numbers of studies, particularly vitamin D (*n* = 75), soy (*n* = 40), *β*-carotene (*n* = 39), folic acid (*n* = 36), calcium (*n* = 27), Mediterranean diet (*n* = 24), and *ω*-3 fatty acids (*n* = 20).

**Figure 9:**
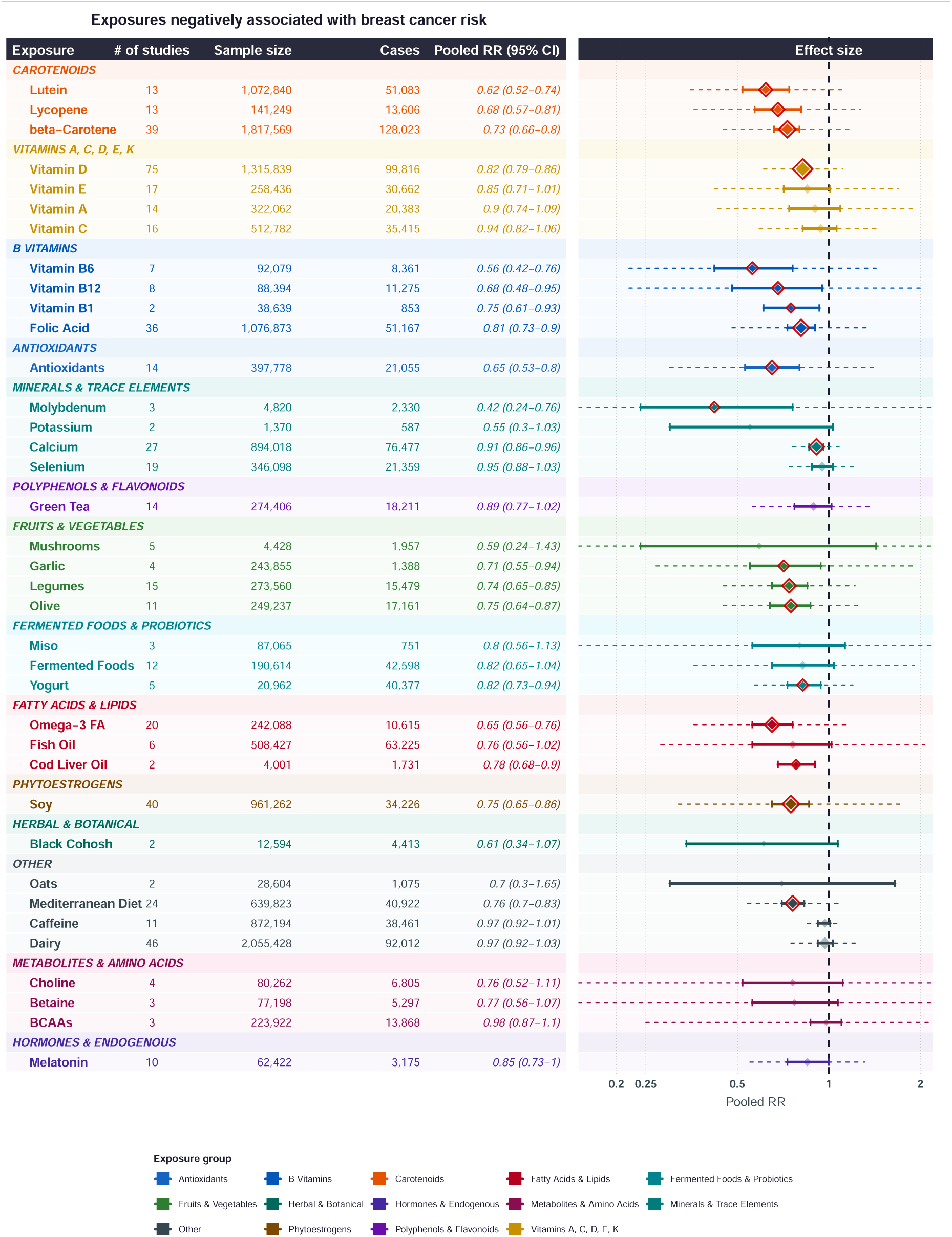
Summary forest plot of nutritional exposures with pooled estimates below 1.00 for breast cancer risk.

After restricting the analysis to dietary-intake studies, significant negative associations remained for vitamin B6, lycopene, lutein, *ω*-3 fatty acids, antioxidants, garlic, folic acid, legumes, soy, vitamin B1, *β*-carotene, Mediterranean diet, cod liver oil, olive, yogurt, vitamin D, and calcium. In addition, mushrooms 0.41 (0.32–0.53), zinc 0.65 (0.49–0.88), and vitamin E 0.81 (0.70–0.94), which were not significantly associated with lower breast cancer risk in the combined analysis, became significantly negatively associated with risk after restriction to dietary-intake studies. In contrast, vitamin B12 0.67 (0.44–1.02) no longer reached statistical significance after the restriction, while molybdenum was not represented in the dietary-intake-only analysis.

To distinguish outcomes of particular interest for follow-up research, we plotted heterogeneity against the pooled effect size (left side of Figure 10). Only exposures evaluated in at least three studies are shown in this plot. A higher position indicates a higher *I*^2^, reflecting a greater proportion of the observed variation in study estimates attributable to between-study heterogeneity rather than sampling variability. Further investigation is warranted to identify potential sources of this heterogeneity, such as differences in study populations, exposure assessment, or dosing, for example using meta-regression. The right side of Figure 10 illustrates the relationship between heterogeneity and the logarithm of the p-value from Egger’s test. Only exposures evaluated in at least 10 studies are shown in this plot. The vertical dashed line represents the conventional significance threshold of *p* = 0.05 for Egger’s test. Exposures appearing to the left of this line have statistically significant Egger’s tests and should be interpreted with additional caution because of potential small-study effects, including publication bias. Although some exposures with high heterogeneity also exhibit small Egger’s-test p-values, this pattern is not consistent, suggesting that small-study effects alone are unlikely to fully explain the observed heterogeneity.

**Figure 10:**
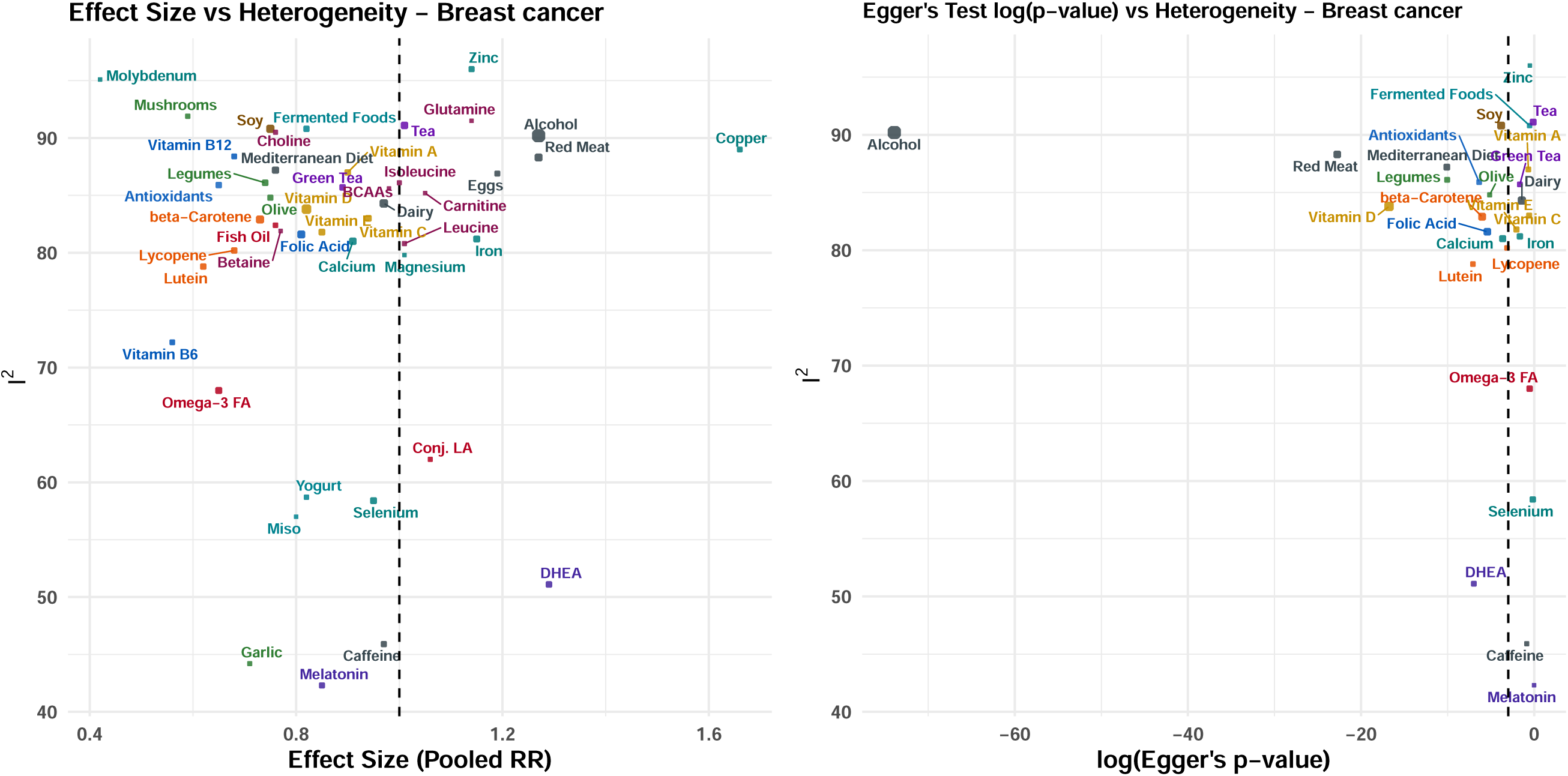
Breast cancer: heterogeneity (*I*^2^) plotted against the pooled RR (left) and the logarithm of the Egger’s-test *p*-value (right).

### 4.2 Exposures associated with ovarian and uterine cancers

Supplemental Figures S1-4 show the summary of exposure effects for ovarian cancer and uterine cancer. Corresponding heterogeneity plots for ovarian and uterine cancers are shown in Supplementary Figures S5 and S6.

For ovarian cancer, among exposures evaluated in at least two studies, no exposure was significantly associated with increased risk. Dairy showed a modest, non-significant positive association, 1.08 (0.98– 1.19). Significant negative associations were observed for selenium 0.21 (0.05–0.92), soy 0.37 (0.22–0.64), antioxidants 0.59 (0.36–0.99), lutein 0.66 (0.45–0.98), vitamin E 0.66 (0.48–0.92), *β*-carotene 0.72 (0.57–0.91), vitamin D 0.72 (0.60–0.87), and *ω*-6 fatty acids 0.78 (0.67–0.90). Other non-significant negative associations were observed for legumes 0.58 (0.22–1.49), vitamin C 0.71 (0.38–1.29), calcium 0.72 (0.48–1.08), folic acid 0.76 (0.56–1.02), tea 0.77 (0.55–1.08), caffeine 0.95 (0.79–1.15), alcohol 0.96 (0.82–1.12), and vitamin A 0.97 (0.86–1.09). When analyses were restricted to dietary intake, soy 0.37 (0.22–0.64), vitamin D 0.52 (0.38–0.71), selenium 0.58 (0.38–0.90), antioxidants 0.59 (0.36–0.99), lutein 0.66 (0.45–0.98), vitamin E 0.66 (0.48–0.92), *β*-carotene 0.72 (0.57–0.91), and *ω*-6 fatty acids 0.78 (0.67–0.90) remained significantly negatively associated with risk. In addition, calcium 0.66 (0.54–0.81), which was not statistically significant in the combined analysis, became significantly negatively associated with ovarian cancer risk after restriction to dietary-intake studies.

For uterine cancer, among exposures evaluated in at least two studies, red meat 1.31 (1.11–1.53), iron 1.52 (1.26–1.82), and copper 2.34 (1.24–4.41) were significantly positively associated with risk. Significant negative associations were observed for folic acid 0.47 (0.31–0.71), lutein 0.47 (0.32–0.69), *β*-carotene 0.57 (0.43–0.76), calcium 0.66 (0.52–0.82), and green tea 0.68 (0.48–0.95). Mediterranean diet, soy, tea, lycopene, vitamin D, *ω*-3 fatty acids, fish oil, caffeine, and alcohol showed non-significant negative associations, with pooled estimates ranging from 0.61 to 0.96. After restricting the analyses to dietary intake, iron 1.53 (1.19– 1.96) and red meat 1.31 (1.11–1.53) remained significantly positively associated with risk, while folic acid 0.47 (0.31–0.71), lutein 0.47 (0.32–0.69), *β*-carotene 0.57 (0.43–0.76), calcium 0.66 (0.52–0.82), and green tea 0.68 (0.48–0.95) remained significantly negatively associated with risk. Copper, although significantly positively associated with uterine cancer risk in the combined analysis, was represented by only one study in the dietary-intake-only analysis and therefore did not meet the two-study threshold.

### 4.3 Comparison of exposure effects across women’s cancers

Figure 11 compares pooled estimates across breast, ovarian, and uterine cancers for shared exposures. Overall, pooled estimates for nutritional exposures more frequently fell below 1.00 than above it, suggesting that negative associations with cancer risk were more common than positive associations across the three cancer types.

**Figure 11:**
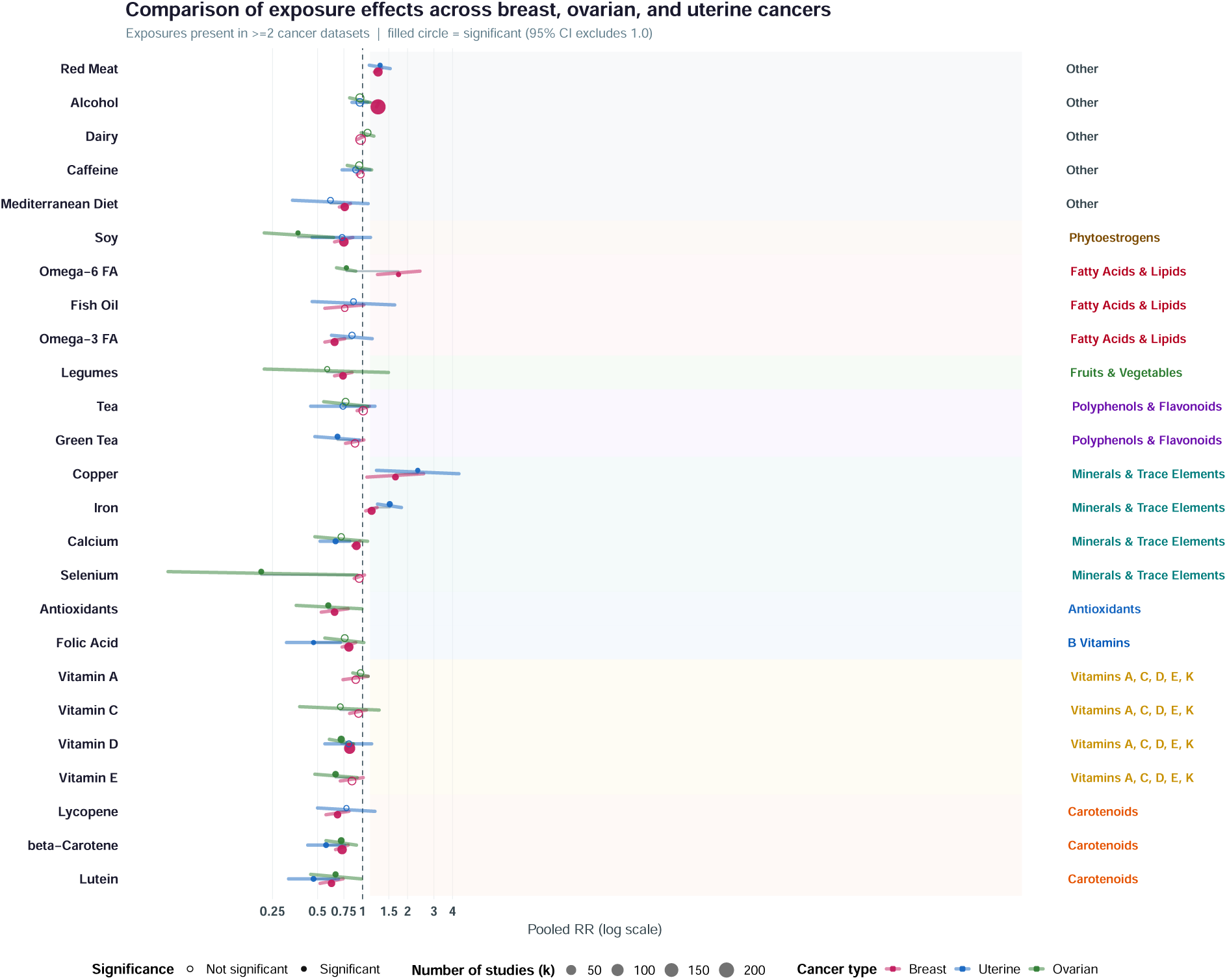
Comparison of pooled relative risks across breast, ovarian, and uterine cancers among exposures evaluated in at least two cancer datasets.

Among exposures associated with lower cancer risk, lutein and *β*-carotene showed the most consistent patterns, with statistically significant negative associations across all three cancer types. For lutein, the pooled estimates were 0.62 (0.52–0.74) for breast, 0.66 (0.45–0.98) for ovarian, and 0.47 (0.32–0.69) for uterine cancer; corresponding estimates for *β*-carotene were 0.73 (0.66–0.80), 0.72 (0.57–0.91), and 0.57 (0.43–0.76), respectively. Several other exposures were significantly associated with lower risk in two cancer types and showed the same direction of association in the third: calcium [breast: 0.91 (0.86–0.96); ovarian: 0.72 (0.48–1.08); uterine: 0.66 (0.52–0.82)], vitamin D [breast: 0.82 (0.79–0.86); ovarian: 0.72 (0.60–0.87); uterine: 0.81 (0.56–1.15)], soy [breast: 0.75 (0.65–0.86); ovarian: 0.37 (0.22–0.64); uterine: 0.73 (0.46–1.13)], and folic acid [breast: 0.81 (0.73–0.90); ovarian: 0.76 (0.56–1.02); uterine: 0.47 (0.31–0.71)]. Antioxidants were also significantly associated with lower risks of both breast cancer [0.65 (0.53–0.80)] and ovarian cancer [0.59 (0.36–0.99)]; the uterine estimate was based on a single study and therefore falls outside the two-study threshold. Vitamin E was significantly associated with lower ovarian cancer risk [0.66 (0.48–0.92)] but did not reach statistical significance for breast cancer in the combined analysis [0.85 (0.71–1.01)]; only one uterine cancer study was available.

The significant negative cross-cancer associations described above remained statistically significant when analyses were restricted to dietary-intake studies. In addition, calcium became significantly associated with lower ovarian cancer risk [0.66 (0.54–0.81)], resulting in significant negative associations with calcium across all three cancer types in the dietary-intake-only analyses [breast: 0.90 (0.86–0.94); ovarian: 0.66 (0.54–0.81); uterine: 0.66 (0.52–0.82)]. Vitamin E, which was not significantly associated with breast cancer risk in the combined analysis, became significantly negatively associated with breast cancer risk after restriction to dietary-intake studies [0.81 (0.70–0.94)] and remained significantly negatively associated with ovarian cancer risk [0.66 (0.48–0.92)].

Among exposures associated with higher cancer risk, iron, red meat, and copper showed significant positive associations with both breast and uterine cancer. Iron was associated with higher risks of breast [1.15 (1.05–1.25)] and uterine cancer [1.52 (1.26–1.82)], while corresponding estimates for red meat were 1.27 (1.20–1.34) and 1.31 (1.11–1.53), respectively. Copper was also significantly positively associated with both breast [1.66 (1.07–2.56)] and uterine cancer [2.34 (1.24–4.41)]. After restricting the analyses to dietary intake, the significant positive associations for iron [breast: 1.22 (1.04–1.43); uterine: 1.53 (1.19–1.96)] and red meat [breast: 1.33 (1.24–1.42); uterine: 1.31 (1.11–1.53)] remained. Copper was represented by only one study in each of the breast and uterine dietary-intake-only analyses and therefore did not meet the two-study threshold.

Among exposures showing conflicting associations across cancer types, *ω*-6 fatty acids demonstrated the clearest pattern, being significantly associated with higher breast cancer risk [1.74 (1.26–2.42)] but lower ovarian cancer risk [0.78 (0.67–0.90)]. These contrasting associations remained statistically significant and were unchanged after restriction to dietary-intake studies [breast: 1.74 (1.26–2.42); ovarian: 0.78 (0.67–0.90)].

Importantly, the volume of evidence was uneven across cancers. Breast cancer generally had the largest number of studies per exposure, while ovarian and uterine cancer estimates were often based on fewer studies and were consequently less precise. Therefore, the cross-cancer comparison should be interpreted primarily as an exploratory analysis rather than as evidence of shared causal effects. Nevertheless, the repeated negative associations observed for several exposures, particularly lutein and *β*-carotene, together with the positive associations observed for iron and red meat and the combined-analysis positive association for copper, suggest patterns that may warrant more targeted investigation across women’s cancers.

### 4.4 Subcategory analysis, multilingual platform and platform update

While most studies have examined breast, uterine, and ovarian cancers as broad disease categories, relatively few have focused on specific cancer subtypes. Supplemental Table S3 summarizes the exposure–subtype associations identified among the included studies. We selected commonly reported subcategories. Because these classifications are not mutually exclusive, some overlap may occur, and a single study may contribute to more than one subcategory. Two exposure–subtype associations are particularly noteworthy. Higher vitamin D exposure was associated with a lower risk of ductal carcinoma in situ (0.79, 0.69–0.89), based on three studies. In contrast, alcohol exposure was associated with a higher risk of invasive lobular carcinoma (1.69, 1.30–2.19), based on five studies included in the meta-analysis.

MetaFemina does not focus on cervical, vulvar, or vaginal cancers because a substantial proportion of these cancers are associated with human papillomavirus (HPV) infection [61]. Socioeconomic status may act as an important unmeasured confounder in studies of nutritional exposures and these cancers. Individuals with greater socioeconomic resources may be more likely to receive HPV vaccination early in life while also having better access to nutritional knowledge and healthier diets. Consequently, differences in HPV vaccination uptake may obscure or distort the observed association between nutritional exposures and cancer risk.

To broaden the accessibility of MetaFemina to researchers worldwide, we provide the platform in multiple languages. The current version supports Simplified Chinese, Traditional Chinese, Dutch, and Korean, in addition to English. The tool also does not restrict articles based on language.. To ensure that MetaFemina incorporates the latest evidence in the field, the platform is updated monthly through GitHub Actions. Relevant exposure data, summary forest plots, and heterogeneity assessment plots displayed on the platform will be updated on the 28th of each month.

## 5 Discussion

Although MetaFemina has demonstrated its efficiency and accuracy compared with existing approaches, our study has its limitations. First, the use of LLMs for automated evidence extraction may introduce errors due to hallucinations or misinterpretation of articles. To mitigate this risk, we implemented a consensus-based approach combining outputs from multiple LLMs and incorporated crowd-sourced verification mechanisms. Second, heterogeneity across studies may arise from differences in exposure measurement, study populations, follow-up periods, and outcome definitions. To account for such variability, we used a random-effects meta-analysis model, which explicitly allows true effect sizes to vary between studies. Third, complex study designs (e.g., longitudinal studies, multi-cohort analyses, or subgroup-specific results) may be difficult to capture reliably through automated extraction alone and may require additional human review. In addition, a single natural food or supplement often contains multiple nutrients that may influence breast cancer risk. For example, garlic is a source of the B vitamins, thiamin and pantothenic acid, as well as the dietary minerals, calcium, potassium, phosphorus, and zinc. As a result, it can be difficult to disentangle the association of any single nutrient independently of the others, since intake of one nutrient is often accompanied by intake of several others. A final limitation is that MetaFemina cannot support causal claims. MetaFemina is designed to support evidence synthesis and hypothesis generation, not to make causal claims. Accordingly, findings reported on the website should be interpreted as associations rather than causal effects. For example, MetaFemina indicates that mushroom dietary intake is associated with a lower breast cancer incidence based on four studies comprising a total of 3,660 participants (pooled RR=0.41, [0.32, 0.53]). Although this association is statistically significant, it does not imply that mushroom intake reduces breast cancer risk. One important concern is unmeasured confounding by socioeconomic status, broader dietary patterns, or health-conscious behavior, and these factors may themselves be associated with breast cancer risk.

Our tool can be readily extended to other exposures and health outcomes. In the future, we intend to expand our platform to study the association between nutritional exposures and cancer prognosis. Public interest in dietary intakes is reflected in a recent study reporting that 89.5% of breast cancer patients used vitamin or mineral supplements and 67.7% used natural products [62]. We also plan to generalize our tool for studying the association between nutritional exposures and other diseases.

## 6 Conclusion

We have developed an LLM-based meta-analysis platform, MetaFemina, for women’s cancers and nutritional exposures. We demonstrate that such an approach can substantially reduce the time and cost compared with traditional evidence synthesis while maintaining accuracy. In addition to pooled estimates, MetaFemina also provides automated assessment of heterogeneity, publication bias, and leave-one-out sensitivity analyses, sample size calculations based on synthesized effect sizes, visual summaries, and plain language interpretations. By enabling trustworthy, reproducible, and up-to-date meta-analyses, this tool has the potential to accelerate meta-analyses and facilitate dissemination of the findings to the fields of oncology and nutritional epidemiology.

## Data Availability

All data produced in the present study are available upon reasonable request to the authors.

https://web-production-c52e1.up.railway.app/

