## Supplemental Materials for "MetaFemina: development and evaluation of a large language model-assisted platform for automated meta-analysis of nutritional exposures and breast, ovarian, and uterine cancer risk"

### Supplementary Material

#### A Technology Stack

The application backend is built in Python using `Flask`, with `pandas` for data manipulation, `NumPy` for numerical operations, `statsmodels` for meta-analytic computations, `Biopython` (`Entrez`) for PubMed access, and `matplotlib` / `forestplot` for visualization. LLM integration uses the `OpenAI API` client (routing to `OpenAI GPT4o`, via the Cornell API proxy) and the `Google GenAI SDK` (`gemini-2.5-flash`). The frontend is implemented in `HTML`, `CSS`, and `JavaScript`. Results are cached in a hierarchical directory structure to avoid redundant API calls.

#### B Literature Search

Relevant studies are identified through automated queries to the PubMed database using the Biopython Entrez API. The user-specified nutritional exposure (e.g., ”Coffee”, ”Vitamin D”, ”Soy”) can be selected from a predefined list derived, in part, from National Institutes of Health resources [1]. Next, the tool constructs a Boolean search query combining the exposure term and the disease term (e.g. breast cancer). Exposure queries are expanded using large language model (LLM)-curated synonym lists (e.g., for ”Soy”: Soybean, Soya, Glycine Max, Tofu, Tempeh, Edamame, Miso, Soy Milk, Soy Protein) to maximize recall. Synonym lists are cached locally to minimize redundant API calls.

Publication type filters are applied depending on user preference: searches by default exclude aggregate evidence (Meta-Analyses, Systematic Reviews). Additional Boolean exclusions are applied to remove animal studies (mice, rat, murine, in vitro), genetic/receptor studies (SNP), unrelated cancer types, and, for incidence analyses, studies focused exclusively on survival among already-diagnosed patients. Full article metadata (title, abstract, authors, journal, publication date, affiliations, publication types) are then fetched in XML format via Entrez.efetch.

As an example, we include the exact PubMed query used for the folic acid and breast cancer incidence analysis:

Listing 1: Example PubMed search for folate and breast cancer risk

```
Searching PubMed for:

(
(Breast AND Cancer)[Title/Abstract]
AND
(
"Folic acid"[Title/Abstract]
OR "folate"[Title/Abstract]
OR "tetrahydrofolate"[Title/Abstract]
OR "5-methyltetrahydrofolate"[Title/Abstract]
OR "serum folate"[Title/Abstract]
OR "plasma folate"[Title/Abstract]
OR "RBC folate"[Title/Abstract]
OR "dietary folate"[Title/Abstract]
OR "folate intake"[Title/Abstract]
OR "folate supplementation"[Title/Abstract]
)
AND
(
incidence
OR risk
OR development
OR "associated with"
OR "odds ratio"
)
NOT
(
"breast cancer survivors"[Title]
OR "breast cancer patients"[Title]
```

```
OR "cancer survivors"[Title]
OR "cancer patients"[Title]
)
NOT
(
mice[Title]
OR mouse[Title]
OR rat[Title]
OR murine[Title]
OR "in vitro"[Title]
)
NOT "SNP"[Title]
NOT
(
polymorphism[Title]
OR polymorphisms[Title]
OR variant[Title]
OR variants[Title]
OR transferase[Title]
OR genotype[Title]
OR genotypes[Title]
OR "telomere length"[Title]
OR "family history"[Title]
OR "gene-diet"[Title]
OR "gene-nutrient"[Title]
OR "gene-supplement"[Title]
OR "genotype-exposure"[Title]
OR "genetic-nutrient"[Title]
)
NOT
(
"interaction"[Title]
AND
(
gene[Title]
OR genetic[Title]
OR polymorphism[Title]
OR mutation[Title]
OR genotype[Title]
)
)
NOT Meta-Analysis[ptyp]
NOT "Systematic Review"[ptyp]
)
```

### C LLM-Based Extraction

Extracted abstracts undergo a two-phase processing pipeline. In Phase 1 (pre-filtering), retrieved articles are screened programmatically for disease relevance, outcome alignment, publication type, and exposure keyword presence. Articles are scored by relevance and sorted accordingly. In Phase 2 (extraction), each abstract is submitted to two LLMs: OpenAI GPT-4o (using the OpenAI API client via proxy) as the primary engine, and Google Gemini 2.5 Flash (using the Google GenAI SDK) serving as both a fallback and validation layer. Each LLM receives a structured prompt instructing it to extract from the abstract: (1) the effect size (HR, OR, or RR) and its 95% confidence interval; (2) sample size and number of cases; (3) study design, timing (prospective/retrospective), geographic continent, and cancer stage; (4) the type of comparison (e.g., highest vs. lowest quartile, per SD increase); (5) a relevance assessment (Relevant, Questionable, or Not Relevant); and (6) a direction-standardisation flag (*needs_inversion*) to ensure all effect sizes are standardised to a high-vs-low exposure reference direction. LLMs return structured JSON responses. Special attention is given to articles reporting inverted effect sizes, for example those examining the impact of deficiency in a given nutrient. Where both LLMs return results, a consensus mechanism is applied specifically for the (*needs_inversion*) flag; inversion of the effect size is only cancelled when both models successfully return results but disagree on the inversion direction. If only one model successfully extracts data, its inversion flag is accepted. A rule-based regex extraction system is employed strictly as a fail-safe fallback only if the LLM screening/extraction pipeline fails to execute due to complete API client or network failures. If the LLM runs successfully and simply determines that no studies meet the inclusion criteria, the tool respects this negative finding and does not trigger the regex fallback, preventing false-positive extractions (e.g., clinical synonyms like "chocolate cysts" for chocolate diet exposures). If neither an effect estimate, a confidence interval, nor a $p$-value specifically associated with an exposure is reported, the article is excluded for that exposure. If an article receives a Fair JBI rating and its full text is available through PMCID, we use ChatGPT 5.6 Luna to re-extract key evidence from the full text.

### D Statistical Analysis

**Random-Effects Meta-Analysis** MetaFemina computes pooled effect sizes using a DerSimonian–Laird random-effects meta-analysis model [2], implemented through the `combine_effects()` function in the `statsmodels` Python library. By default, studies with fewer than 50 cases are excluded from the pooled analysis.

Let $\hat{\mu}_i$ denote the effect estimate (log relative risk) from study $i$, and let $v_i$ denote its within-study sampling variance, for $i = 1, \ldots, K$. Under a random-effects meta-analysis model,

$$\hat{\mu}_i \mid \mu_i \sim N(\mu_i, v_i),$$

where $\mu_i$ is the true effect in study $i$. The study-specific true effects are assumed to vary around an overall mean effect $\mu$:

$$\mu_i \sim N(\mu, \tau^2),$$

where $\tau^2$ is the between-study variance. Marginally, this gives

$$\hat{\mu}_i \sim N(\mu, v_i + \tau^2).$$

The inverse-variance weight assigned to study $i$ is therefore

$$w_i = \frac{1}{v_i + \tau^2}. \tag{1}$$

After estimating $\tau^2$, the pooled random-effects estimate is

$$\hat{\mu} = \frac{\sum_{i=1}^{K} w_i \hat{\mu}_i}{\sum_{i=1}^{K} w_i}. \tag{2}$$

Thus, studies with smaller within-study variance $v_i$ receive greater weight, but the between-study variance $\tau^2$ reduces the dominance of any single large study when heterogeneity is present.

**Heterogeneity Assessment** Between-study heterogeneity is commonly quantified using the statistics $I^2$ and $\tau^2$, which are derived from the random-effects meta-analysis model. The parameter $\tau^2$ represents the estimated between-study variance and reflects the extent to which the true treatment effects differ across studies. In contrast, the $I^2$ statistic describes the proportion of the total variability in observed study estimates that is attributable to true heterogeneity rather than sampling error [3]. Values of $I^2$ range from 0% to 100%, where larger values indicate greater heterogeneity between studies. These measures provide complementary information about the magnitude of between-study variation and are automatically reported in our meta-analyses.

**Publication Bias** Potential publication bias is assessed using funnel plots and Egger's regression test for funnel plot asymmetry [4]. Let $s_i = \sqrt{v_i}$ denote the standard error of the study-specific effect estimate $\hat{\mu}_i$. Egger's test is based on the regression

$$\frac{\hat{\mu}_i}{s_i} = \beta_0 + \beta_1 \frac{1}{s_i} + \varepsilon_i, \qquad i = 1, \ldots, I, \tag{3}$$

where $\hat{\mu}_i/s_i$ is the standardized effect estimate and $1/s_i$ is the study precision. Evidence against $H_0 : \beta_0 = 0$ suggests funnel plot asymmetry and potential small-study or publication bias.

For visualization, funnel plots display the study-specific log effect estimates $\hat{\mu}_i$ against their standard errors $s_i$. The pooled random-effects estimate $\hat{\mu}$ from (2) is shown as the center line, and the pseudo-95% confidence limits, $\hat{\mu}\pm 1.96 s_i$, are shown as two dashed lines forming the funnel shape. Because standard errors generally decrease with sample size, $s_i = \sqrt{v_i} \propto 1/\sqrt{n_i}$, larger studies are expected to provide more precise effect estimates and therefore cluster near the pooled estimate $\hat{\mu}$. Smaller studies have larger standard errors and are expected to scatter more widely around $\hat{\mu}$, approximately within the pseudo-95% limits $\hat{\mu} \pm 1.96 s_i$. In the absence of small-study effects or publication bias, this scatter should be roughly symmetric around the pooled estimate; marked asymmetry may indicate that studies on one side of the funnel are missing or underrepresented.

**Sensitivity Analyses** A leave-one-out analysis is performed, iteratively re-running the meta-analysis excluding each study in turn to assess the robustness of the pooled estimate to the inclusion of any single study. A Baujat plot [5] is generated to visualize each study's contribution to the overall heterogeneity (Cochran's $Q$ statistic) against its influence on the pooled effect estimate. Thus, studies located toward the right side of the plot contribute more strongly to between-study heterogeneity, whereas studies located higher on the plot have a greater impact on the summary estimate.

For study $i$, the contribution to heterogeneity can be expressed as

$$Q_i = w_i(\hat{\mu}_i - \hat{\mu})^2,$$

where $\hat{\mu}_i$ is the study-specific effect estimate, $w_i$ is the corresponding inverse-variance weight, and $\hat{\mu}$ is the pooled effect estimate.

In MetaFemina, the Baujat plot is provided as a graphical sensitivity analysis rather than a formal hypothesis test. Studies appearing in the upper-right region of the plot are flagged for closer review because they have both large heterogeneity contributions and large influence on the pooled estimate. This information is interpreted together with leave-one-out analyses, funnel plots, Egger's regression test, study quality scores, and study-level design characteristics. A study identified as influential by the Baujat plot is not automatically excluded from the meta-analysis; instead, the plot provides users with additional evidence to assess the robustness of the pooled result and to determine whether sensitivity analyses excluding influential studies are warranted.

**Visualization** Forest plots are generated using the `forestplot` Python library, displaying individual study log-transformed effect sizes with 95% confidence intervals. Plot dimensions and font sizes scale dynamically based on the number of included studies. Funnel plots and Baujat plots are generated using `matplotlib`.

### E Supplementary Table

Table S1: Additional studies on folate-related exposures and breast cancer risk identified by MetaFemina but not included in Eleotério et al. [6]

| Authors | Location | Sample size/Cases | Design | Exposure assessment | Exposure groups | Effect estimate (95% CI) |
|---|---|---|---|---|---|---|
| Freudenheim et al. [7] | USA | 608/297 | Retrospective case–control | FFQ; dietary folate and related nutrients | $>$ 460 versus $<$ 304 $\mu$g/day of dietary folate | OR 0.50 (0.31–0.82) |
| Sellers et al. [8] | USA | 34,387/1,586 | Prospective cohort | FFQ; dietary folate | $>$ 294 versus $\leq$ 172 $\mu$g/day of dietary folate; estimate reoriented from the reported low-versus-high comparison | RR 0.8264 (0.621–1.099) |
| Sellers et al. [9] | USA | 34,393/1,875 | Prospective cohort | FFQ; dietary folate | Higher versus lower folate intake; estimate inverted to high versus low | RR 0.6993 (0.495–0.9804) |
| Zhang et al. [10] | USA | 32,826/712 | Prospective nested case–control | Prediagnostic plasma folate | $>$ 14.0 versus $<$ 4.6 ng/mL plasma folate | RR 0.73 (0.50–1.07) |
| Adzersen et al. [11] | Germany | 663/310 | Retrospective case–control | FFQ; dietary folate and selected micronutrients | $>$ 147 versus $<$ 77 $\mu$g/day of folate equivalents | OR 0.47 (0.25–0.88) |
| Baglietto et al. [12] | Australia | 17,447/537 | Prospective cohort | FFQ; dietary folate | 200 versus 400 $\mu$g/day of dietary folate | HR 0.77 (0.33–1.80) |
| Zhang et al. [13] | USA | 88,744/3,797 | Prospective cohort | Repeated FFQs; total folate | $\geq$ 534 versus $\leq$ 228 $\mu$g/day of total folate | RR 0.81 (0.66–0.99) |
| Stolzenberg-Solomon et al. [14] | USA | 25,400/691 | Prospective cohort | Baseline dietary questionnaire; total folate from food and supplements | $>$ 853.0 versus $\leq$ 335.5 $\mu$g/day of total folate | HR 1.32 (1.04–1.68) |
| Lajous et al. [15] | Mexico | 1,866/475 | Retrospective case–control | FFQ; dietary folate, vitamin B6, and vitamin B12 | Highest versus lowest category; median dietary folate 454 versus 224 $\mu$g/day | OR 0.64 (0.45–0.90) |
| Lin et al. [16] | USA | 28,345/848 | Prospective nested case–control | Prediagnostic plasma folate | $>$ 15.8 versus $\leq$ 5.1 ng/mL plasma folate | RR 1.42 (1.00–2.02) |
| Zhang et al. [17] | USA | 5,442/379 | Randomized controlled trial | Daily intervention: folic acid 2.5 mg, vitamin B6 50 mg, and vitamin B12 1 mg | Active combined B-vitamin treatment versus placebo | HR 0.83 (0.60–1.14) |
| Ericson et al. [18] | Sweden | 612/204 | Prospective nested case–control | Prediagnostic plasma folate | Highest versus lowest tertile; median plasma folate 17 versus 6 nmol/L | OR 2.67 (1.44–4.92) |
| Stevens et al. [19] | USA | 70,656/3,898 | Prospective cohort | FFQ; dietary folate | $\geq$ 312.1 versus $<$ 166.9 $\mu$g/day of dietary folate | HR 1.12 (1.01–1.24) |
| Zhang et al. [20] | China | 876/438 | Retrospective case–control | FFQ; dietary folate and one-carbon nutrients | Highest versus lowest study-specific dietary-folate category | OR 0.32 (0.21–0.49) |
| Shrubsole et al. [21] | China | 72,861/718 | Prospective cohort | FFQ; dietary folate and other B vitamins | Highest versus lowest quintile; mean dietary folate 404 versus 194 $\mu$g/day | HR 0.58 (0.34–0.99) |
| Islam et al. [22] | Japan | 5,262/1,754 | Retrospective case–control | FFQ; dietary folate | Highest versus lowest tertile of dietary folate | OR 0.79 (0.68–0.93) |
| Yang et al. [23] | USA | 4,850/2,325 | Retrospective case–control | FFQ; dietary folate, B vitamins, and methionine | $>$ 893 versus $<$ 405 $\mu$g/day of dietary folate | OR 0.50 (0.25–1.00) |
| Gong et al. [24] | USA | 3,016/1,493 | Retrospective case–control | FFQ; natural-food folate | Q4 $>$ 315.1 versus Q1 $\leq$ 159.6 $\mu$g/day of natural-food folate | OR 0.57 (0.33–1.00) |
| Rukundo et al. [25] | Uganda | 145/72 | Retrospective case–control | RBC folate biomarker | Normal versus low RBC folate | OR 1.40 (0.70–2.90) |
| Cancarini et al. [26] | Italy | 10,786/391 | Prospective cohort | FFQ; dietary folate | Highest versus lowest quartile of dietary folate | RR 0.65 (0.44–0.95) |
| de Batlle et al. [27] | Europe | 367,993/11,575 | Prospective EPIC cohort | Country-specific FFQs; dietary folate | Highest versus lowest quintile (Q5 versus Q1) | HR 0.92 (0.83–1.01) |
| Essén et al. [28] | Sweden | 19,775/795 | Prospective cohort | Baseline serum folate | High $>$ 32 versus normal 5–32 nmol/L among fasting participants | HR 1.47 (1.06–2.04) |
| Houghton et al. [29] | USA | 3,748/1,874 | Prospective nested case–control | Prediagnostic plasma folate and one-carbon metabolites | Highest versus lowest category for plasma collected before mandatory folic-acid fortification | RR 0.93 (0.75–1.16) |
| Houghton et al. [30] | USA | 1,817/610 | Prospective nested case–control | Prediagnostic plasma folate and one-carbon metabolites | Highest versus lowest study-specific plasma-folate category | RR 1.18 (0.84–1.66) |

Continued on next page

Table S1 continued

| Authors | Location | Sample size/Cases | Design | Exposure assessment | Exposure groups | Effect estimate (95% CI) |
|---|---|---|---|---|---|---|
| Sánchez-Zamorano et al. [31] | Mexico | 636/342 | Retrospective case–control | FFQ-derived folate intake | Highest versus lowest folate intake; estimate inverted to high versus low | OR 0.1089 (0.0304–0.3906) |
| Hatami et al. [32] | Iran | 305/151 | Retrospective case–control | FFQ; total folate and other one-carbon vitamins | Q4 $\geq$ 309.57 versus Q1 $\leq$ 188.87 $\mu$g/day of total folate | OR 0.09 (0.04–0.21) |
| Koenig et al. [33] | USA | 1,612/553 | Prospective nested case–control | Prediagnostic serum UMFA and storage-corrected 5-MTHF | Highest versus lowest quintile of storage-corrected 5-MTHF | OR 0.69 (0.49–0.97) |

*Note:* Light-blue rows indicate records whose exposure measurement was not classified as `dietary intake`, including human-biospecimen, unclear, or mixed exposures.
*Abbreviations:* 5-MTHF, 5-methyltetrahydrofolate; FFQ, food-frequency questionnaire; MTHFR, methylenetetrahydrofolate reductase; Q, quantile; RBC, red blood cell; UMFA, unmetabolized folic acid.

Table S2: Additional studies on vitamin E exposures and breast cancer risk identified by MetaFemina but not included in de Oliveira et al. [34]

| Authors | Location | Sample size/Cases | Design | Exposure assessment | Exposure groups | Effect estimate (95% CI) |
|---|---|---|---|---|---|---|
| London et al. [35] | USA | 953/377 | Retrospective case–control | Dietary questionnaire; vitamin E intake from food sources | Highest vs Lowest | OR 0.40 (0.20–0.90) |
| Lee et al. [36] | USA | 39,876/NR | Randomized controlled trial | Natural-source $\alpha$-tocopherol supplementation | Vitamin E supplementation vs Placebo | RR 1.00 (0.90–1.12) |
| Hunter et al. [37] | USA | 89,494/1,439 | Prospective cohort | Semiquantitative FFQ; vitamins C, E, and A intake | Highest vs Lowest | RR 0.99 (0.83–1.19) |
| Maillard et al. [38] | France | 19,934/366 | Prospective case–control | Serum $\alpha$-tocopherol and related biomarkers | Highest vs Lowest | OR 0.68 (0.41–1.10) |
| Nagel et al. [39] | Europe | 345,995/7,502 | Prospective cohort | Country-specific dietary questionnaires; dietary vitamin E | Highest vs Lowest | HR 1.11 (0.84–1.46) |
| Frazier et al. [40] | USA | 47,355/361 | Retrospective cohort | High-school FFQ; adolescent dietary vitamin E | Q5 vs Q1 | RR 0.61 (0.42–0.89) |
| Playdon et al. [41] | USA | 1,242/621 | Prospective case–control | Nutritional metabolomics; vitamin E-related metabolites | 90th percentile vs 10th percentile | OR 1.64 (1.18–2.28) |
| Fernández-Lázaro et al. [42] | Spain | 9,983/107 | Prospective cohort | Validated semiquantitative FFQ; dietary antioxidant vitamins and minerals | T3 vs T1 (Highest vs Lowest) | HR 0.92 (0.55–1.54) |
| Gerber et al. [43] | Italy/France | 663/319 | Retrospective case–control | Biological assays; liposoluble vitamin status | Highest vs Lowest | OR 4.20 (1.90–9.00) |
| van 't Veer et al. [44] | Europe | 721/347 | Retrospective case–control | Adipose-tissue antioxidant biomarkers | Highest vs Lowest | OR 1.15 (0.75–1.77) |
| Freudenheim et al. [7] | USA | 608/297 | Retrospective case–control | FFQ; dietary vitamin E and related nutrients | Highest vs Lowest | OR 0.55 (0.34–0.88) |
| Sharhar et al. [45] | Asia | 196/57 | Retrospective case–control | Plasma vitamin E concentration | Low plasma vitamin E ($<$2.5 mg/dl) vs Higher levels (displayed as high vs low) | OR 0.3367 (0.2874–0.7246) |
| Bonilla-Fernández et al. [46] | Mexico | 282/141 | Retrospective case–control | Dietary questionnaire; nutritional factors including vitamin E | Highest vs Lowest | OR 0.10 (0.02–0.44) |

*Note:* Light-blue rows indicate records whose exposure measurement was not classified as `dietary intake`, including human-biospecimen, unclear, or mixed exposures. *Abbreviations:* COMT, catechol-O-methyltransferase; FFQ, food-frequency questionnaire; NR, not reported; Q, quantile; T, tertile.

Table S3: Meta-analysis results for exposures evaluated in breast, ovarian, and uterine cancer subcategories. Only exposure–subcategory pairs with at least two included studies are shown. Statistically significant associations are boldfaced.

| Subcategory | Exposure | Studies identified/Studies included | Pooled RR (95% CI) |
|---|---|---|---|
| **Breast cancer** | | | |
| Ductal carcinoma in situ | **Vitamin D** | **3/3** | **0.79 (0.69–0.89)** |
| Invasive ductal carcinoma | Alcohol | 4/4 | 1.28 (0.96–1.70) |
| Invasive lobular carcinoma | **Alcohol** | **5/5** | **1.69 (1.30–2.19)** |
| Triple-negative breast cancer | Alcohol | 6/6 | 1.11 (0.80–1.52) |
| | Vitamin D | 4/4 | 0.99 (0.41–2.42) |
| **Ovarian cancer** | | | |
| Mucinous carcinoma | Alcohol | 3/3 | 1.73 (0.88–3.43) |
| **Uterine cancer** | | | |
| *No exposure–subcategory pair had at least two included studies.* | | | |

### F Supplemental Figures

#### F.1 Forest plots for ovarian and uterine cancers

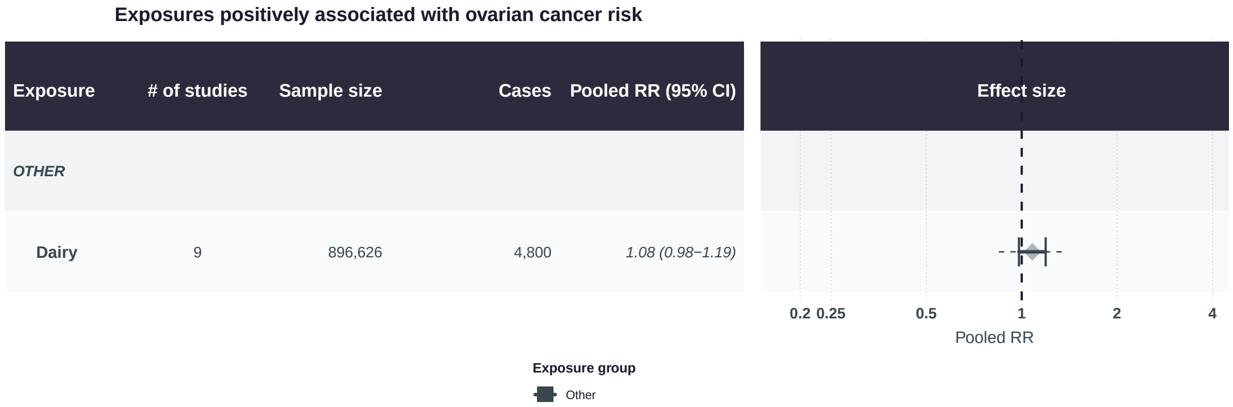


Figure S1: Summary forest plot of nutritional exposure with pooled estimates above 1.00 for ovarian cancer risk.

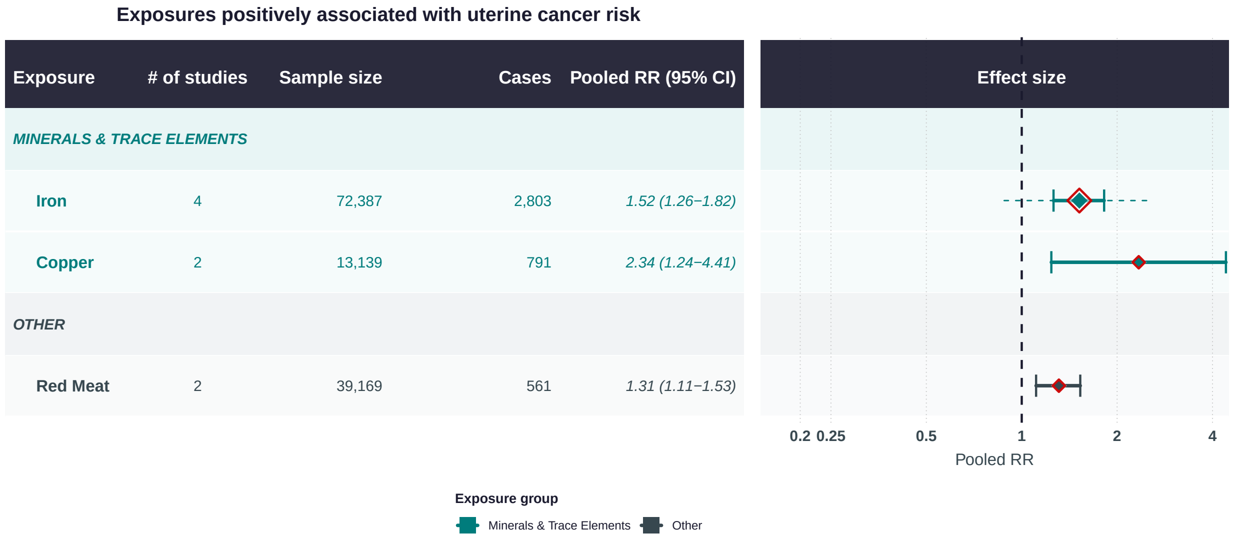


Figure S2: Summary forest plot of nutritional exposures with pooled estimates above 1.00 for uterine cancer risk.

**Exposures negatively associated with ovarian cancer risk**

| Exposure | # of studies | Sample size | Cases | Pooled RR (95% CI) |
|---|---|---|---|---|
| *CAROTENOIDS* | | | | |
| **Lutein** | 4 | 529,715 | 3,624 | *0.66 (0.45–0.98)* |
| **beta–Carotene** | 8 | 611,872 | 4,692 | *0.72 (0.57–0.91)* |
| *VITAMINS A, C, D, E, K* | | | | |
| **Vitamin E** | 6 | 591,029 | 4,497 | *0.66 (0.48–0.92)* |
| **Vitamin C** | 3 | 583,089 | 2,528 | *0.71 (0.38–1.29)* |
| **Vitamin D** | 11 | 68,361 | 5,262 | *0.72 (0.6–0.87)* |
| **Vitamin A** | 6 | 1,588,466 | 9,351 | *0.97 (0.86–1.09)* |
| *B VITAMINS* | | | | |
| **Folic Acid** | 8 | 723,200 | 3,495 | *0.76 (0.56–1.02)* |
| *ANTIOXIDANTS* | | | | |
| **Antioxidants** | 4 | 523,871 | 2,791 | *0.59 (0.36–0.99)* |
| *MINERALS & TRACE ELEMENTS* | | | | |
| **Selenium** | 3 | 1,947 | 768 | *0.21 (0.05–0.92)* |
| **Calcium** | 5 | 37,261 | 2,041 | *0.72 (0.48–1.08)* |
| *POLYPHENOLS & FLAVONOIDS* | | | | |
| **Tea** | 8 | 307,213 | 4,002 | *0.77 (0.55–1.08)* |
| *FRUITS & VEGETABLES* | | | | |
| **Legumes** | 2 | 63,479 | 506 | *0.58 (0.22–1.49)* |
| *FATTY ACIDS & LIPIDS* | | | | |
| **Omega−6 FA** | 2 | 6,630 | 3,238 | *0.78 (0.67–0.9)* |
| *PHYTOESTROGENS* | | | | |
| **Soy** | 2 | 1,906 | 754 | *0.37 (0.22–0.64)* |
| *OTHER* | | | | |
| **Caffeine** | 11 | 336,054 | 38,479 | *0.95 (0.79–1.15)* |
| **Alcohol** | 20 | 832,445 | 18,046 | *0.96 (0.82–1.12)* |

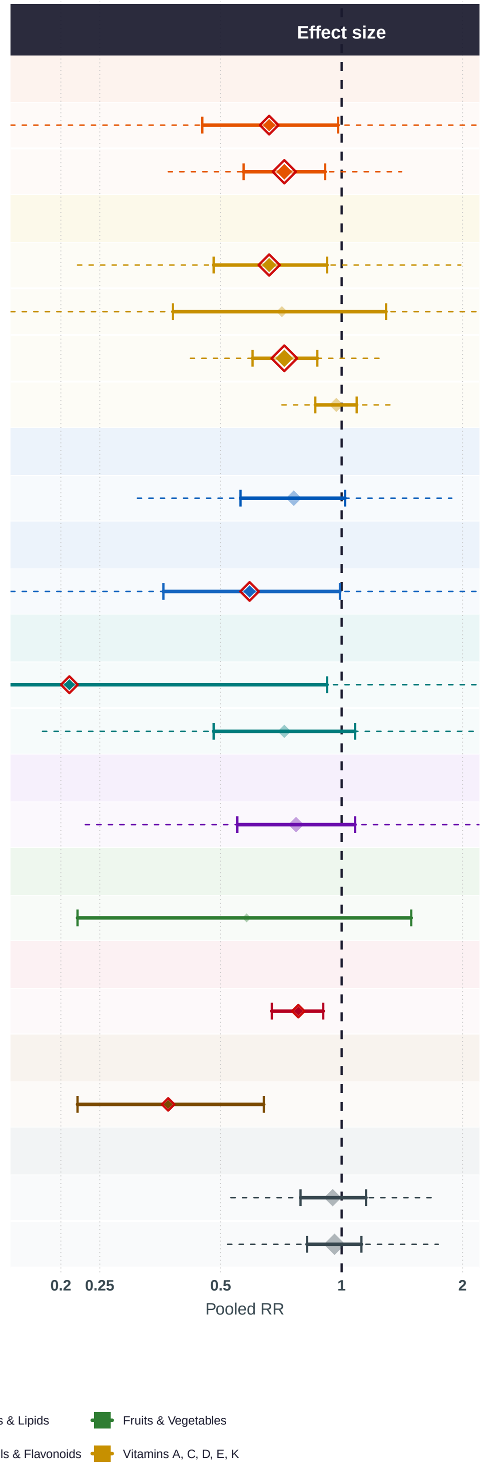


Figure S3: Summary forest plot of nutritional exposures with pooled estimates below 1.00 for ovarian cancer risk.

**Exposures negatively associated with uterine cancer risk**

| Exposure | # of studies | Sample size | Cases | Pooled RR (95% CI) |
|---|---|---|---|---|
| *CAROTENOIDS* | | | | |
| Lutein | 3 | 3,315 | 1,227 | *0.47 (0.32–0.69)* |
| beta–Carotene | 3 | 3,315 | 1,227 | *0.57 (0.43–0.76)* |
| Lycopene | 2 | 2,233 | 686 | *0.78 (0.5–1.21)* |
| *VITAMINS A, C, D, E, K* | | | | |
| Vitamin D | 4 | 66,026 | 2,398 | *0.81 (0.56–1.15)* |
| *B VITAMINS* | | | | |
| Folic Acid | 2 | 74,680 | 412 | *0.47 (0.31–0.71)* |
| *MINERALS & TRACE ELEMENTS* | | | | |
| Calcium | 4 | 6,240 | 1,445 | *0.66 (0.52–0.82)* |
| *POLYPHENOLS & FLAVONOIDS* | | | | |
| Green Tea | 3 | 3,331 | 1,564 | *0.68 (0.48–0.95)* |
| Tea | 4 | 4,413 | 2,105 | *0.74 (0.45–1.21)* |
| *FATTY ACIDS & LIPIDS* | | | | |
| Omega–3 FA | 5 | 457,863 | 4,240 | *0.85 (0.62–1.16)* |
| Fish Oil | 3 | 110,943 | 2,072 | *0.87 (0.46–1.64)* |
| *PHYTOESTROGENS* | | | | |
| Soy | 3 | 95,991 | 933 | *0.73 (0.46–1.13)* |
| *OTHER* | | | | |
| Mediterranean Diet | 3 | 90,098 | 3,100 | *0.61 (0.34–1.09)* |
| Caffeine | 4 | 150,854 | 1,222 | *0.9 (0.73–1.12)* |
| Alcohol | 15 | 411,289 | 8,229 | *0.96 (0.85–1.08)* |

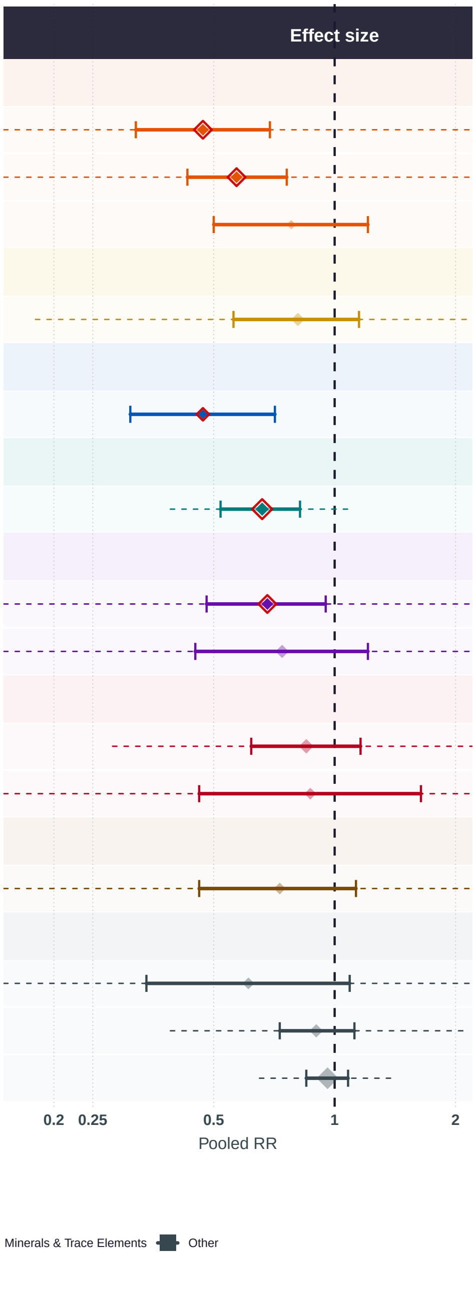


Figure S4: Summary forest plot of nutritional exposures with pooled estimates below 1.00 for uterine cancer risk.

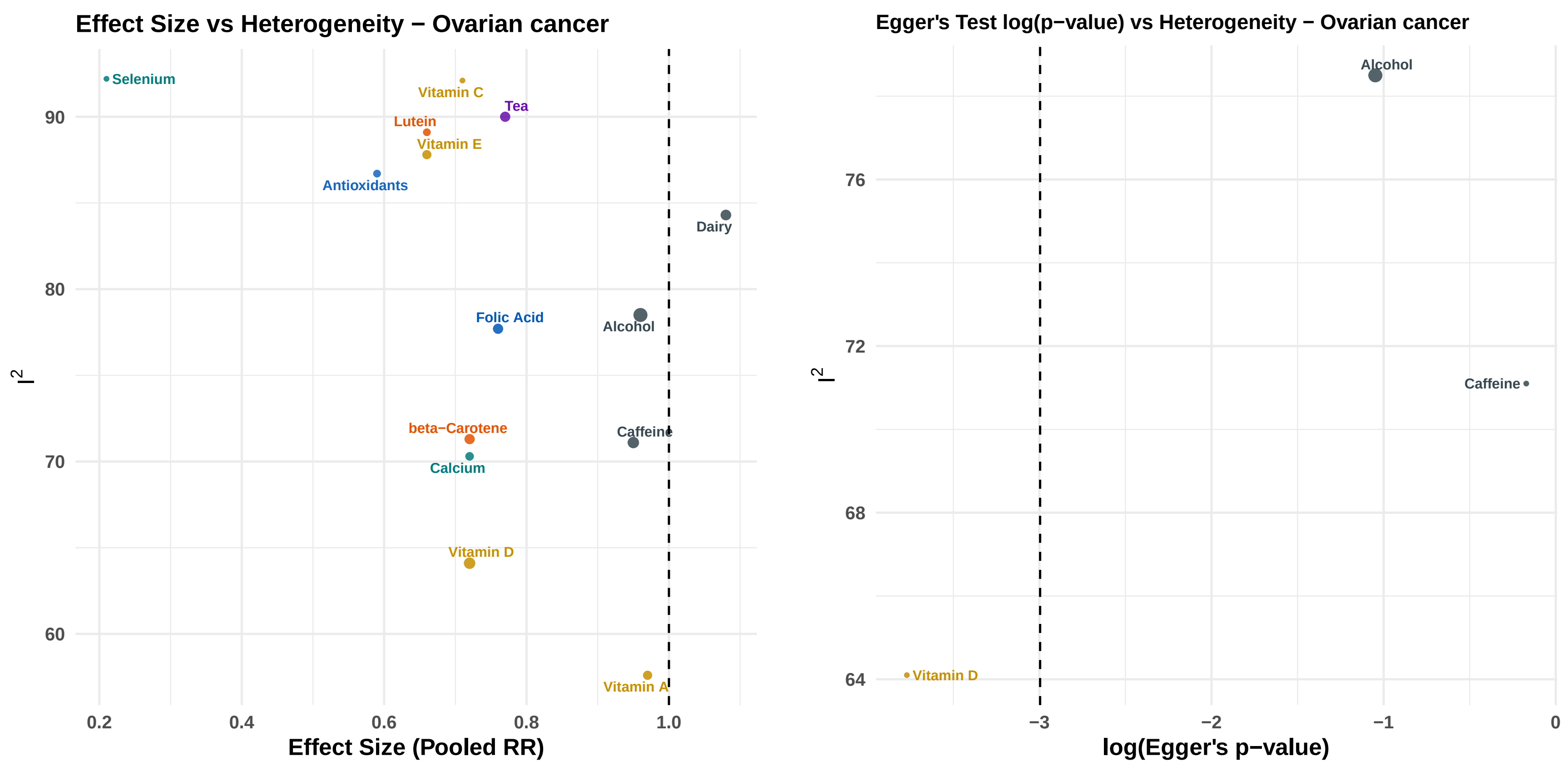


Figure S5: Ovarian cancer: heterogeneity ($I^2$) plotted against the pooled RR (left) and the logarithm of the Egger's-test $p$-value (right).

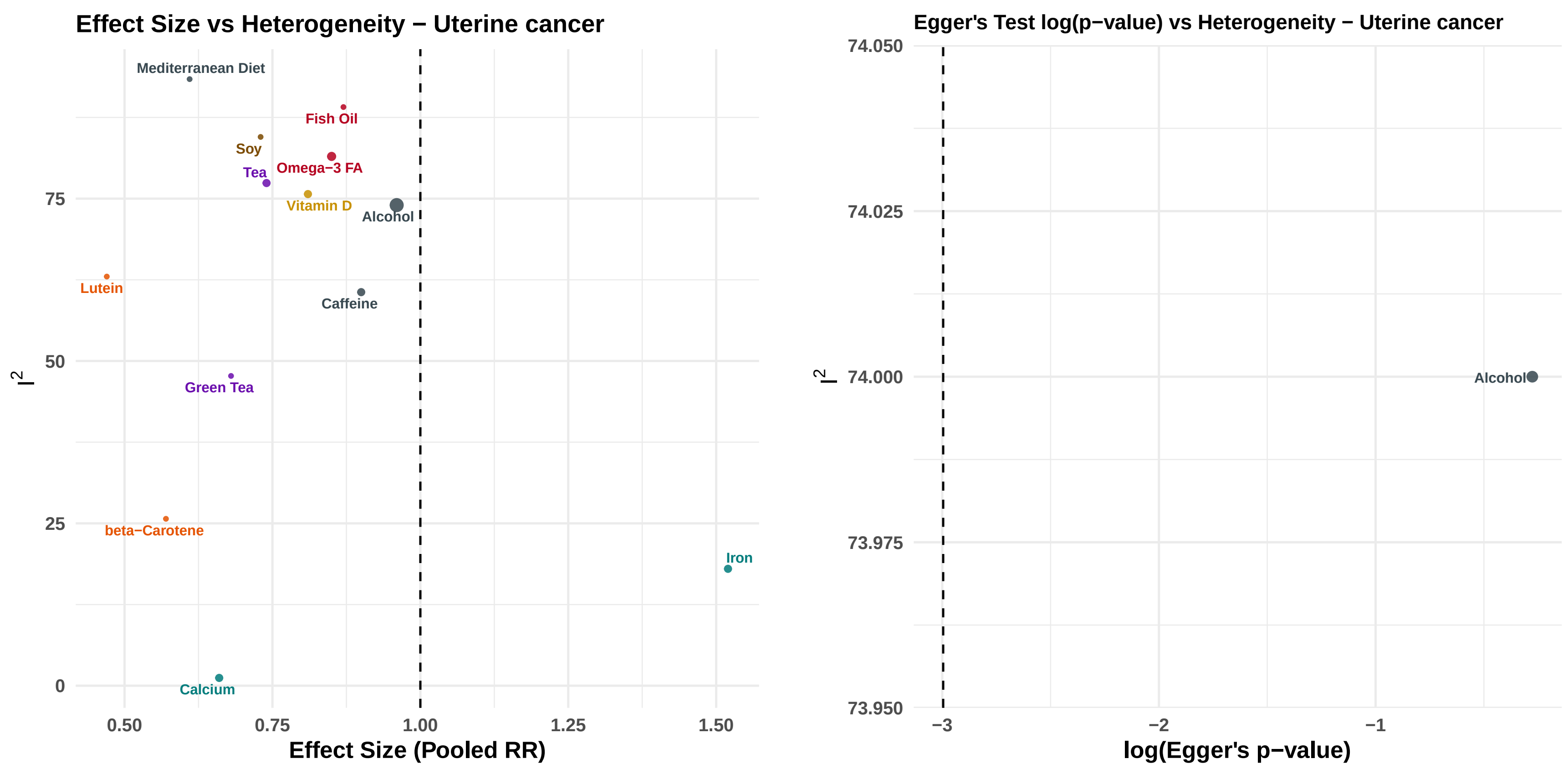


Figure S6: Uterine cancer: heterogeneity ($I^2$) plotted against the pooled RR (left) and the logarithm of the Egger's-test $p$-value (right).

#### F.2 Forest plots comparing exposures for dietary intake studies

**Exposures positively associated with breast cancer risk
meta−analysis of dietary−intake studies**

| Exposure | # of studies | Sample size | Cases | Pooled RR (95% CI) |
|---|---|---|---|---|
| *VITAMINS A, C, D, E, K* | | | | |
| Vitamin K | 2 | 5,048 | 2,592 | *1.01 (0.68−1.5)* |
| *MINERALS & TRACE ELEMENTS* | | | | |
| Selenium | 6 | 175,826 | 12,809 | *1.02 (0.87−1.2)* |
| Iron | 8 | 385,425 | 22,415 | *1.22 (1.04−1.43)* |
| *POLYPHENOLS & FLAVONOIDS* | | | | |
| Tea | 24 | 1,181,650 | 65,433 | *1.02 (0.93−1.13)* |
| *FRUITS & VEGETABLES* | | | | |
| Grapefruit | 2 | 164,504 | 5,404 | *1.1 (0.8−1.52)* |
| *FATTY ACIDS & LIPIDS* | | | | |
| Conj. LA | 4 | 380,443 | 17,329 | *1.06 (0.92−1.22)* |
| Omega−6 FA | 2 | 2,476 | 1,201 | *1.74 (1.26−2.42)* |
| *OTHER* | | | | |
| Eggs | 8 | 29,311 | 5,421 | *1.19 (0.99−1.43)* |
| Alcohol | 182 | 16,035,924 | 1,275,429 | *1.24 (1.22−1.27)* |
| Red Meat | 34 | 1,178,141 | 60,075 | *1.33 (1.24−1.42)* |

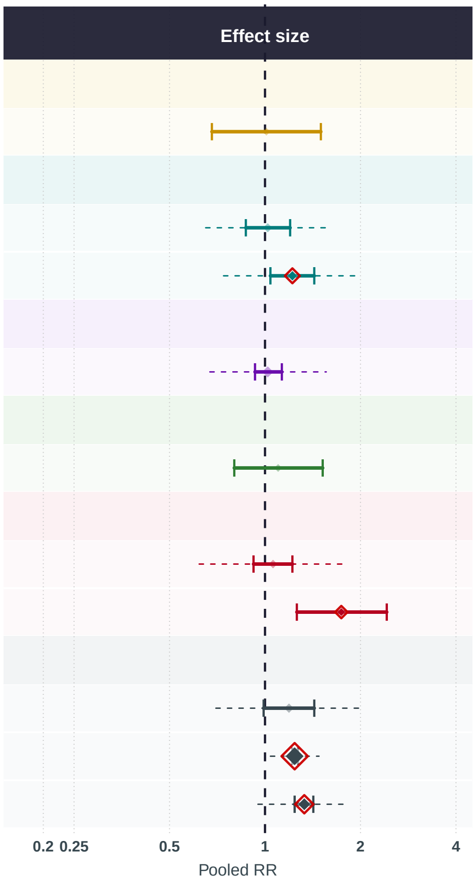


**Exposure group**
Fatty Acids & Lipids · Fruits & Vegetables · Minerals & Trace Elements · Other · Polyphenols & Flavonoids

Figure S7: Summary forest plot of nutritional exposures with pooled estimates above 1.00 for breast cancer risk based on dietary intake studies.

**Exposures negatively associated with breast cancer risk**
**meta−analysis of dietary−intake studies**

| Exposure | # of studies | Sample size | Cases | Pooled RR (95% CI) |
|---|---|---|---|---|
| *CAROTENOIDS* | | | | |
| Lycopene | 5 | 90,844 | 5,625 | 0.55 (0.36−0.84) |
| Lutein | 7 | 1,061,834 | 45,572 | 0.66 (0.55−0.8) |
| beta−Carotene | 25 | 1,684,061 | 114,896 | 0.77 (0.69−0.85) |
| *VITAMINS A, C, D, E, K* | | | | |
| Vitamin E | 12 | 235,680 | 28,952 | 0.81 (0.7−0.94) |
| Vitamin D | 24 | 1,048,907 | 55,763 | 0.85 (0.79−0.92) |
| Vitamin A | 8 | 287,201 | 16,059 | 0.89 (0.66−1.19) |
| Vitamin C | 16 | 512,782 | 35,415 | 0.94 (0.82−1.06) |
| *B VITAMINS* | | | | |
| Vitamin B6 | 4 | 52,399 | 6,236 | 0.4 (0.24−0.67) |
| Vitamin B12 | 5 | 38,106 | 7,877 | 0.67 (0.44−1.02) |
| Folic Acid | 28 | 987,993 | 45,499 | 0.74 (0.66−0.83) |
| Vitamin B1 | 2 | 38,639 | 853 | 0.75 (0.61−0.93) |
| *ANTIOXIDANTS* | | | | |
| Antioxidants | 9 | 392,499 | 18,643 | 0.7 (0.55−0.88) |
| *MINERALS & TRACE ELEMENTS* | | | | |
| Potassium | 2 | 1,370 | 587 | 0.55 (0.3−1.03) |
| Zinc | 5 | 43,927 | 7,652 | 0.65 (0.49−0.88) |
| Calcium | 20 | 871,662 | 74,218 | 0.9 (0.86−0.94) |
| Magnesium | 2 | 69,324 | 9,766 | 0.91 (0.79−1.06) |
| *POLYPHENOLS & FLAVONOIDS* | | | | |
| Green Tea | 13 | 250,180 | 18,067 | 0.89 (0.77−1.02) |
| *FRUITS & VEGETABLES* | | | | |
| Mushrooms | 4 | 3,660 | 1,829 | 0.41 (0.32−0.53) |
| Garlic | 4 | 243,855 | 1,388 | 0.71 (0.55−0.94) |
| Legumes | 15 | 273,560 | 15,479 | 0.74 (0.65−0.85) |
| Olive | 10 | 248,595 | 16,870 | 0.8 (0.69−0.92) |
| *FERMENTED FOODS & PROBIOTICS* | | | | |
| Miso | 3 | 87,065 | 751 | 0.8 (0.56−1.13) |
| Fermented Foods | 12 | 190,614 | 42,598 | 0.82 (0.65−1.04) |
| Yogurt | 5 | 20,962 | 40,377 | 0.82 (0.73−0.94) |
| *FATTY ACIDS & LIPIDS* | | | | |
| Omega−3 FA | 13 | 238,479 | 7,644 | 0.67 (0.57−0.8) |
| Fish Oil | 4 | 508,253 | 62,658 | 0.74 (0.49−1.11) |
| Cod Liver Oil | 2 | 4,001 | 1,731 | 0.78 (0.68−0.9) |
| *PHYTOESTROGENS* | | | | |
| Soy | 35 | 919,838 | 32,658 | 0.75 (0.64−0.87) |
| *HERBAL & BOTANICAL* | | | | |
| Black Cohosh | 2 | 12,594 | 4,413 | 0.61 (0.34−1.07) |
| *OTHER* | | | | |
| Oats | 2 | 28,604 | 1,075 | 0.7 (0.3−1.65) |
| Mediterranean Diet | 24 | 639,823 | 40,922 | 0.76 (0.7−0.83) |
| Caffeine | 11 | 872,194 | 38,461 | 0.97 (0.92−1.01) |
| Dairy | 45 | 2,055,428 | 91,218 | 0.97 (0.92−1.02) |
| *METABOLITES & AMINO ACIDS* | | | | |
| Choline | 3 | 79,262 | 6,305 | 0.74 (0.45−1.21) |
| BCAAs | 2 | 197,211 | 10,396 | 0.77 (0.4−1.48) |
| Isoleucine | 2 | 197,211 | 10,396 | 0.77 (0.4−1.48) |
| Leucine | 2 | 197,211 | 10,396 | 0.77 (0.4−1.48) |
| Betaine | 2 | 76,198 | 4,797 | 0.79 (0.5−1.25) |

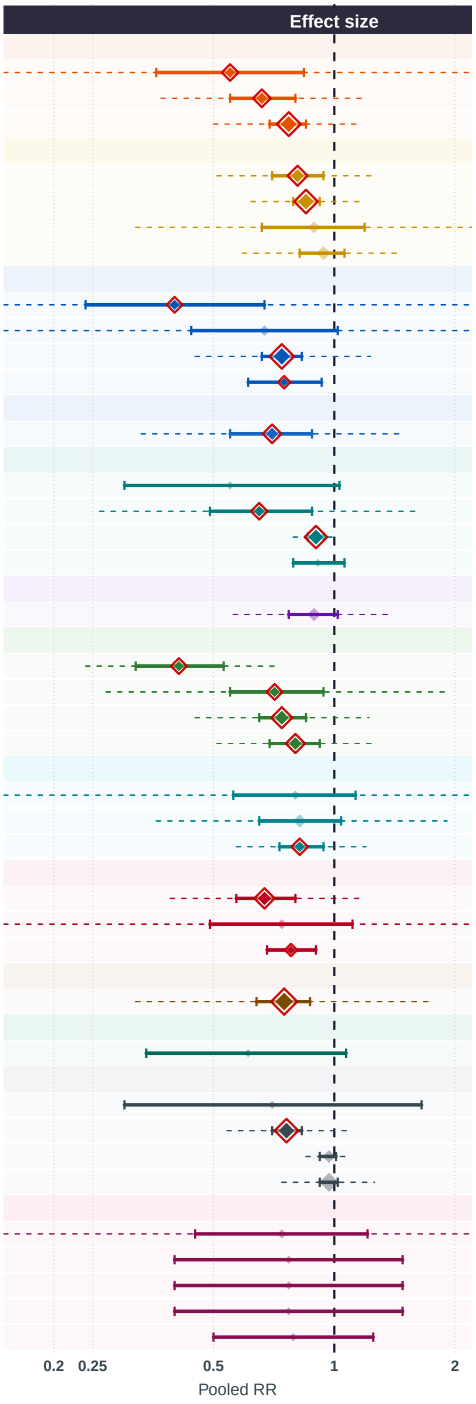


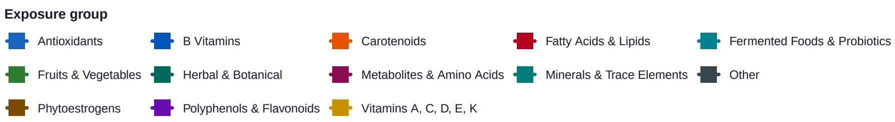


Figure S8: Summary forest plot of nutritional exposures with pooled estimates below 1.00 for breast cancer risk based on dietary intake studies.

**Exposures negatively associated with ovarian cancer risk**
**meta−analysis of dietary−intake studies**

| Exposure | # of studies | Sample size | Cases | Pooled RR (95% CI) |
|---|---|---|---|---|
| *CAROTENOIDS* | | | | |
| **Lutein** | 4 | 529,715 | 3,624 | *0.66 (0.45−0.98)* |
| **beta−Carotene** | 8 | 611,872 | 4,692 | *0.72 (0.57−0.91)* |
| *VITAMINS A, C, D, E, K* | | | | |
| **Vitamin D** | 2 | 4,611 | 1,993 | *0.52 (0.38−0.71)* |
| **Vitamin E** | 6 | 591,029 | 4,497 | *0.66 (0.48−0.92)* |
| **Vitamin C** | 3 | 583,089 | 2,528 | *0.71 (0.38−1.29)* |
| **Vitamin A** | 6 | 1,588,466 | 9,351 | *0.97 (0.86−1.09)* |
| *B VITAMINS* | | | | |
| **Folic Acid** | 8 | 723,200 | 3,495 | *0.76 (0.56−1.02)* |
| *ANTIOXIDANTS* | | | | |
| **Antioxidants** | 4 | 523,871 | 2,791 | *0.59 (0.36−0.99)* |
| *MINERALS & TRACE ELEMENTS* | | | | |
| **Selenium** | 2 | 1,633 | 611 | *0.58 (0.38−0.9)* |
| **Calcium** | 3 | 36,513 | 1,667 | *0.66 (0.54−0.81)* |
| *POLYPHENOLS & FLAVONOIDS* | | | | |
| **Tea** | 8 | 307,213 | 4,002 | *0.77 (0.55−1.08)* |
| *FRUITS & VEGETABLES* | | | | |
| **Legumes** | 2 | 63,479 | 506 | *0.58 (0.22−1.49)* |
| *FATTY ACIDS & LIPIDS* | | | | |
| **Omega−6 FA** | 2 | 6,630 | 3,238 | *0.78 (0.67−0.9)* |
| *PHYTOESTROGENS* | | | | |
| **Soy** | 2 | 1,906 | 754 | *0.37 (0.22−0.64)* |
| *OTHER* | | | | |
| **Alcohol** | 18 | 819,111 | 17,098 | *0.95 (0.82−1.11)* |
| **Caffeine** | 11 | 336,054 | 38,479 | *0.95 (0.79−1.15)* |

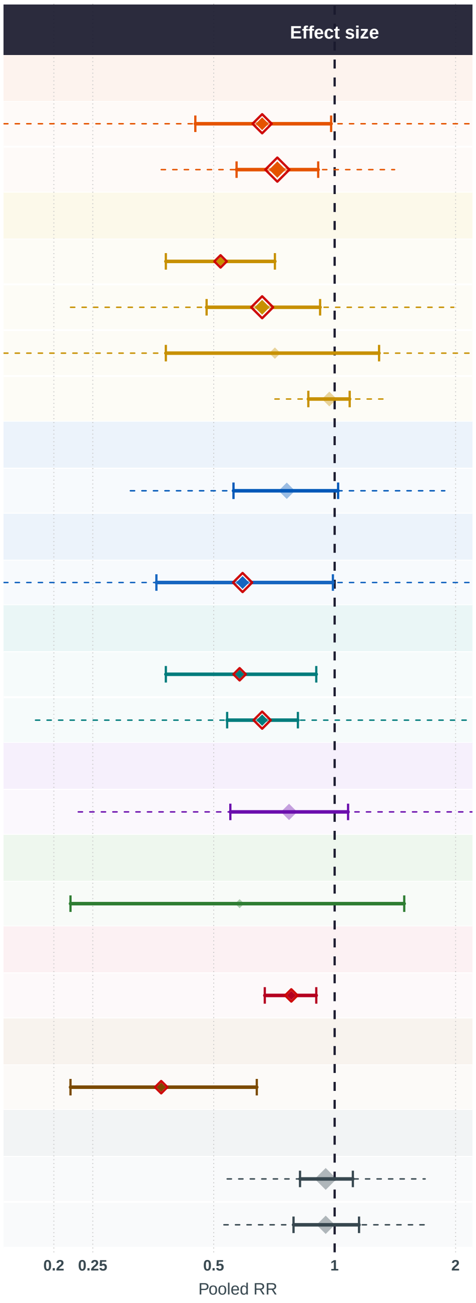


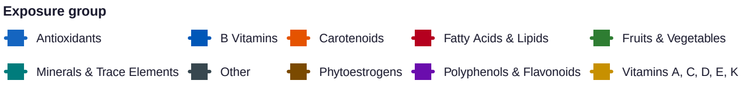


Figure S9: Summary forest plot of nutritional exposures with pooled estimates below 1.00 for ovarian cancer risk based on dietary intake studies.

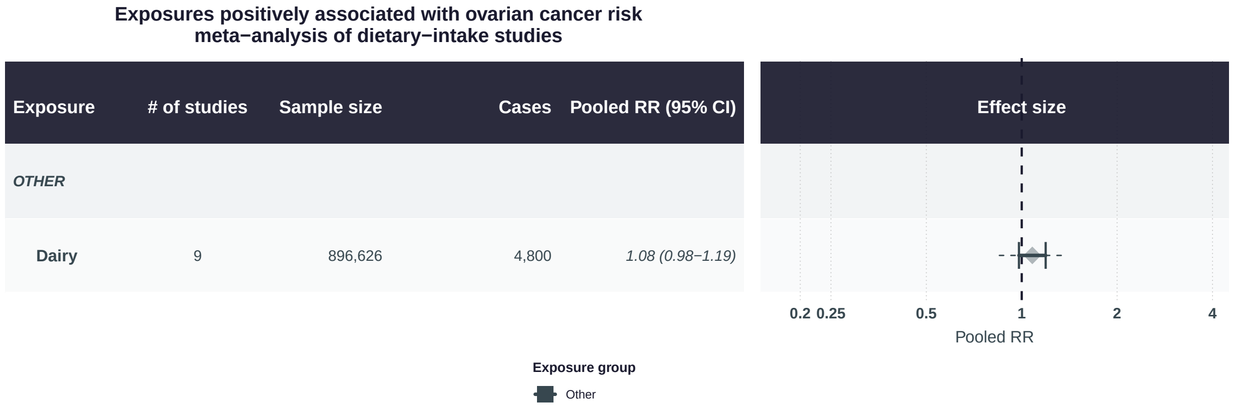


Figure S10: Summary forest plot of nutritional exposure with pooled estimates above 1.00 for ovarian cancer risk based on dietary intake studies.

**Exposures negatively associated with uterine cancer risk**
**meta−analysis of dietary−intake studies**

| Exposure | # of studies | Sample size | Cases | Pooled RR (95% CI) |
|---|---|---|---|---|
| *CAROTENOIDS* | | | | |
| Lutein | 3 | 3,315 | 1,227 | *0.47 (0.32−0.69)* |
| beta−Carotene | 3 | 3,315 | 1,227 | *0.57 (0.43−0.76)* |
| Lycopene | 2 | 2,233 | 686 | *0.78 (0.5−1.21)* |
| *B VITAMINS* | | | | |
| Folic Acid | 2 | 74,680 | 412 | *0.47 (0.31−0.71)* |
| *MINERALS & TRACE ELEMENTS* | | | | |
| Calcium | 4 | 6,240 | 1,445 | *0.66 (0.52−0.82)* |
| *POLYPHENOLS & FLAVONOIDS* | | | | |
| Green Tea | 3 | 3,331 | 1,564 | *0.68 (0.48−0.95)* |
| Tea | 4 | 4,413 | 2,105 | *0.74 (0.45−1.21)* |
| *FATTY ACIDS & LIPIDS* | | | | |
| Omega−3 FA | 5 | 457,863 | 4,240 | *0.85 (0.62−1.16)* |
| Fish Oil | 3 | 110,943 | 2,072 | *0.87 (0.46−1.64)* |
| *PHYTOESTROGENS* | | | | |
| Soy | 3 | 95,991 | 933 | *0.73 (0.46−1.13)* |
| *OTHER* | | | | |
| Mediterranean Diet | 3 | 90,098 | 3,100 | *0.61 (0.34−1.09)* |
| Caffeine | 4 | 150,854 | 1,222 | *0.9 (0.73−1.12)* |
| Alcohol | 15 | 411,289 | 8,229 | *0.96 (0.85−1.08)* |

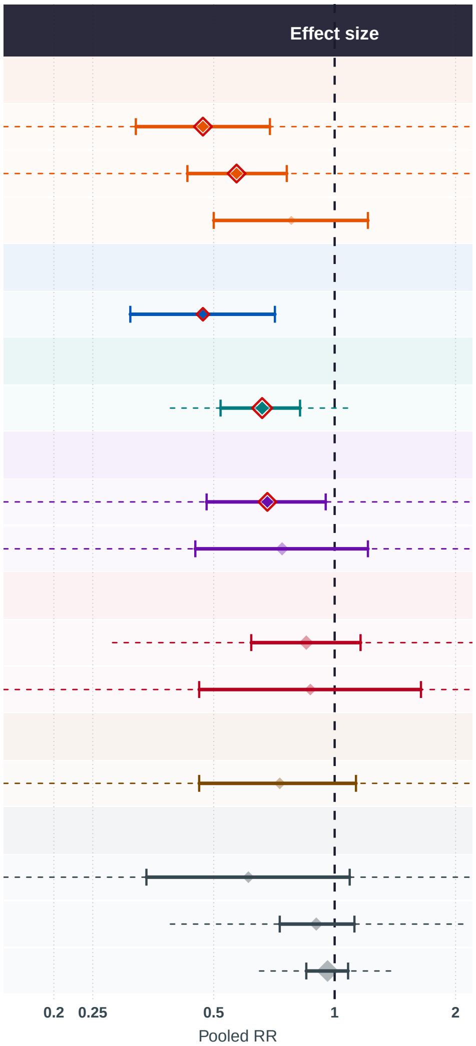


**Exposure group**
B Vitamins · Carotenoids · Fatty Acids & Lipids · Minerals & Trace Elements · Other · Phytoestrogens · Polyphenols & Flavonoids

Figure S11: Summary forest plot of nutritional exposures with pooled estimates below 1.00 for uterine cancer risk based on dietary intake studies.

**Exposures positively associated with uterine cancer risk
meta−analysis of dietary−intake studies**

| Exposure | # of studies | Sample size | Cases | Pooled RR (95% CI) |
|---|---|---|---|---|
| *MINERALS & TRACE ELEMENTS* | | | | |
| Iron | 3 | 66,907 | 2,633 | *1.53 (1.19−1.96)* |
| *OTHER* | | | | |
| Red Meat | 2 | 39,169 | 561 | *1.31 (1.11−1.53)* |

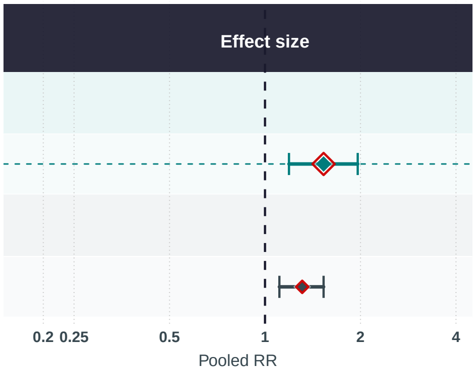


**Exposure group**

Minerals & Trace Elements  Other

Figure S12: Summary forest plot of nutritional exposures with pooled estimates above 1.00 for uterine cancer risk based on dietary intake studies.

[27] De Batlle JD, Ferrari P, Chajes V, Park JY, Slimani N, McKenzie F, Overvad K, Roswall N, Tjønneland A, Boutron-Ruault MC, Clavel-Chapelon F, Fagherazzi G, Katzke V, Kaaks R, Bergmann MM, Trichopoulou A, Lagiou P, Trichopoulos D, Palli D, Sieri S, Panico S, Tumino R, Vineis P, Bueno-de-Mesquita HB, Peeters PH, Hjartåker A, Engeset D, Weiderpass E, Sánchez S, Travier N, Sánchez MJ, Amiano P, Chirlaque MD, Gurrea AB, Khaw KT, Key TJ, Bradbury KE, Ericson U, Sonestedt E, Van Guelpen B, Schneede J, Riboli E, Romieu I. Dietary folate intake and breast cancer risk: European Prospective Investigation into Cancer and Nutrition. Journal of the National Cancer Institute 2015; 107:dju367. DOI: 10.1093/jnci/dju367

[39] Nagel G, Linseisen J, van Gils CH, Peeters PH, Boutron-Ruault MC, Clavel-Chapelon F, Romieu I, Tjønneland A, Olsen A, Roswall N, Witt PM, Overvad K, Rohrmann S, Kaaks R, Drogan D, Boeing H, Trichopoulou A, Stratigakou V, Zylis D, Engeset D, Lund E, Skeie G, Berrino F, Grioni S, Mattiello A, Masala G, Tumino R, Zanetti R, Ros MM, Bueno-de-Mesquita HB, Ardanaz E, Sánchez MJ, Huerta JM, Amiano P, Rodríguez L, Manjer J, Wirfält E, Lenner P, Hallmans G, Spencer EA, Key TJ, Bingham S, Khaw KT, Rinaldi S, Slimani N, Boffetta P, Gallo V, Norat T, Riboli E. Dietary beta-carotene, vitamin C and E intake and breast cancer risk in the European Prospective Investigation into Cancer and Nutrition (EPIC). Breast Cancer Research and Treatment 2010; 119:753–65. DOI: 10.1007/s10549-009-0444-8
